# Acute Respiratory Distress Syndrome is associated with impaired alveolar macrophage efferocytosis

**DOI:** 10.1101/2021.03.15.21253591

**Authors:** Rahul Y. Mahida, Aaron Scott, Dhruv Parekh, Sebastian T. Lugg, Rowan S. Hardy, Gareth G. Lavery, Michael A. Matthay, Babu Naidu, Gavin D. Perkins, David R. Thickett

**Affiliations:** Birmingham Acute Care Research Group, Institute of Inflammation and Ageing, University of Birmingham, Birmingham, U.K; Institute of Metabolism and Systems Research, University of Birmingham, Birmingham, U.K; Cardiovascular Research Institute, Department of Medicine, and Department of Anaesthesia, University of California San Francisco, San Francisco, California, U.S.A; Emergency, Pre-hospital, Perioperative and Critical Care Group, Warwick Medical School, University of Warwick, Warwick, U.K

## Abstract

**Background:** Alveolar macrophage dysfunction may contribute to Acute Respiratory Distress Syndrome (ARDS) pathogenesis, however this has been little studied. Objective: To investigate the pathophysiological link between alveolar macrophage efferocytosis, alveolar neutrophil apoptosis and clinical outcomes in ARDS patients, and to determine whether efferocytosis can be restored.

**Methods:** Ventilated sepsis patients with or without ARDS underwent broncho-alveolar lavage. Apoptosis of alveolar neutrophils was assessed using flow cytometry. Alveolar macrophages were isolated and used in flow cytometric efferocytosis assays with labelled apoptotic neutrophils. Alveolar macrophages were also isolated from the lung tissue of lobectomy patients, then treated with pooled ARDS BAL fluid prior to functional assessment. Rac1 gene expression was assessed using RT-qPCR.

**Results:** Patients with sepsis-related ARDS have decreased alveolar macrophage efferocytosis and increased alveolar neutrophil apoptosis compared to control ventilated sepsis patients. Across all ventilated sepsis patients, alveolar macrophage efferocytosis correlated negatively with alveolar cytokines (IL-8, IL-1ra), duration of ventilation and mortality. ARDS BAL treatment of alveolar macrophages decreased efferocytosis and Rac1 gene expression, however bacterial phagocytosis was preserved. Unexpectedly, alveolar macrophage efferocytosis receptor expression (MerTK, CD206) decreased and expression of the anti-efferocytosis receptor SIRPα increased following ARDS BAL treatment. Rho-associated kinase inhibition partially restored alveolar macrophage efferocytosis in an *in vitro* model of ARDS.

**Conclusions:** Patients with sepsis-related ARDS have impaired AM efferocytosis, which potentially contributes to ARDS pathogenesis and negatively impacts clinical outcomes, including mortality. Strategies to upregulate AM efferocytosis may be of value for attenuating inflammation in ARDS.

## INTRODUCTION

Acute respiratory distress syndrome (ARDS) is an inflammatory disorder of the lungs, with sepsis being the underlying aetiology in >75% of cases(1). Despite advances in supportive care and ventilation strategies, mortality for moderate to severe ARDS remains at 40-46%, and treatment options are limited(1). We now understand more about how ARDS develops: it requires damage to the alveolar epithelium and endothelium(2), leading to reduced alveolar fluid clearance(3), increased permeability, exaggerated inflammation, and a neutrophilic alveolar oedema(4). However, the role of alveolar macrophages (AMs) in ARDS pathogenesis is not fully understood.

ARDS is associated with neutrophil influx into alveoli. Persistently high neutrophil and low AM numbers in broncho-alveolar lavage (BAL) fluid are associated with greater mortality(5). Murine models of ARDS have shown that large numbers of neutrophils undergo apoptosis and accumulate within alveoli(6). Therefore, while the inflammatory alveolar environment of early ARDS may initially delay apoptosis, these neutrophils ultimately undergo apoptosis within alveoli. Efficient efferocytosis of these apoptotic neutrophils by AMs is critical for the resolution of inflammation in ARDS and promotion of tissue repair(6, 7). Apoptotic neutrophils may accumulate in ARDS due to defective AM efferocytosis and/or overwhelmed AM efferocytosis capacity from excessive apoptosis(8). These apoptotic neutrophils then undergo secondary necrosis, resulting in cell rupture and release of inflammatory contents (i.e. Damage Associated Molecular Patterns) into surrounding tissues(9). These mediators are postulated to cause prolonged inflammation, which is characteristic of the ARDS lung microenvironment(10).

AM expression of the Mer tyrosine kinase (MerTK) receptor may be critical for efferocytosis(11). MerTK signalling via phosphatidylinositol 3’-OH kinase (PI3K) results in activation of Rac1, which causes cytoskeletal re-arrangement and engulfment of the apoptotic cell(12, 13). AM surface receptors including CD206 and CD163 are also thought to mediate efferocytosis(14). AMs also express signal regulatory protein-α (SIRPα) on their surface, which binds surfactant proteins or CD47 on healthy cells(15). SIRPα signalling activates Rho-associated kinase (ROCK), and phosphatase and tensin homologue (PTEN) which oppose PI3K signalling, resulting in Rac1 inhibition and suppression of efferocytosis(16). The impact of ARDS on AM expression of these surface receptors and intracellular pathways is unknown.

No study has previously assessed AM efferocytosis in ARDS patients. However monocyte-derived macrophages (MDMs) from ARDS patients do have impaired efferocytosis, and treatment of healthy MDMs with ARDS patient BAL suppresses efferocytosis(17). We hypothesized that AMs in ARDS have impaired efferocytosis, resulting in persistent inflammation due to secondary neutrophil necrosis, which influences clinical outcome. This study had the following aims:

1. To determine if sepsis patients with ARDS have a greater impairment in AM efferocytosis and elevated alveolar neutrophil apoptosis compared to sepsis patients without ARDS
2. To determine if there is an association between AM efferocytosis, alveolar inflammation and clinical outcomes in ventilated sepsis patients with, or without, ARDS.
3. To determine if an *in vitro* model can replicate the impaired efferocytosis observed in ARDS AMs, and thus be used to investigate the mechanism of this impairment.

Some results of these studies have been previously reported in an abstract(18).

## METHODS

Detailed methods are available in the online supplement.

### Ethical Approval

Ethical approval was obtained to recruit ventilated sepsis patients with and without ARDS (REC 16/WA/0169), and for the use of lung tissue samples from patients undergoing routine thoracic surgery (REC 17/WM/0272). For patients who lacked capacity, permission to enrol was sought from a personal legal representative in accordance with the UK Mental Capacity Act (2005). For patients with capacity, written informed consent was obtained from the patient.

### Patient recruitment and broncho-alveolar lavage

Invasively ventilated adult patients with sepsis were recruited from the intensive care unit of the Queen Elizabeth Hospital Birmingham, U.K. from December 2016 – February 2019. Sepsis was defined according to Sepsis-3 criteria(19). Patients who fulfilled the Berlin criteria for ARDS(20) within the previous 48 hours were classified as the ARDS group, whereas those without ARDS were defined as the ventilated control group. Exclusion criteria included imminent treatment withdrawal, steroid therapy prior to admission, abnormal clotting function precluding bronchoscopy, clinically relevant immunosuppression for any reason, or if assent could not be obtained. Recruited patients underwent bronchoscopy with broncho-alveolar lavage within 48 hours of initiation of mechanical ventilation. Sterile, isotonic saline (2 × 50mls) was instilled into a sub-segmental bronchus of the lingula or middle lobe. Recovery ranged between 15 – 60 ml and did not differ between patient groups. BAL fluid was immediately filtered and placed on ice.

### Alveolar Macrophage isolation and functional assessment

Lung tissue samples were obtained from never-smoker or long-term ex-smoker (quit >5 years) adult patients, with normal spirometry and no history of asthma or chronic obstructive pulmonary disease (COPD), all undergoing lobectomy for malignancy at Birmingham Heartlands Hospital, U.K. from 2017-2019. No patient received chemotherapy prior to surgery. Non-affected lung tissue samples underwent saline lavage.

AMs were isolated from patient BAL and lung tissue lavage fluid using Lymphoprep density gradient centrifugation and plastic adherence(21). After 24 hours, AMs were used in efferocytosis and phagocytosis assays. Lung tissue resection AMs were treated with pooled acellular ARDS BAL to create an *in vitro* model of ARDS. AM surface receptor expression and gene expression were assessed. BAL neutrophil apoptosis was also assessed. See supplemental methods for full details.

### Statistical Analysis

Data were analysed using Prism 8 software (GraphPad, USA). Parametric data shown as mean and standard deviation. Non-parametric data shown as median and interquartile range. Differences between continuously distributed data assessed using t-tests for parametric data, or Mann-Whitney tests for non-parametric data. Differences between non-parametric paired data assessed using Wilcoxon matched-pairs signed rank test. Differences between three or more unpaired parametric data sets assessed using Analysis of Variance (ANOVA) followed by Dunn’s multiple comparison tests. Differences between three or more paired parametric data sets assessed using the repeated measures ANOVA followed by Tukey’s multiple comparison tests. Log-rank test used for survival curve comparison. Two-tailed p-values of ≤0.05 were considered significant.

## RESULTS

### Patient characteristics

A total of 38 ventilated sepsis patients were recruited; 21 of which had ARDS, the remainder being controls (table 1). Of the control patients, 4 developed ARDS later in their admission. BAL was collected from 31 patients (17 ARDS, 14 control). Of these, BAL neutrophil apoptosis was assessed in 21 patients (12 ARDS, 9 control). Neutrophil apoptosis was not initially assessed and was added to the study protocol after recruitment had begun. Of those patients who had BAL collected, AM yield was sufficient to perform efferocytosis in 22 patients (11 ARDS, 11 control); mean AM yield was 1.2 million cells. The flow diagram with details of the number of patients included in each assay is in supplemental figure 2. BAL cytokine content from both patient groups was quantified (supplemental table 1). AMs were also isolated from the lung tissue of 16 patients who underwent lobectomy.

**Table 1:**
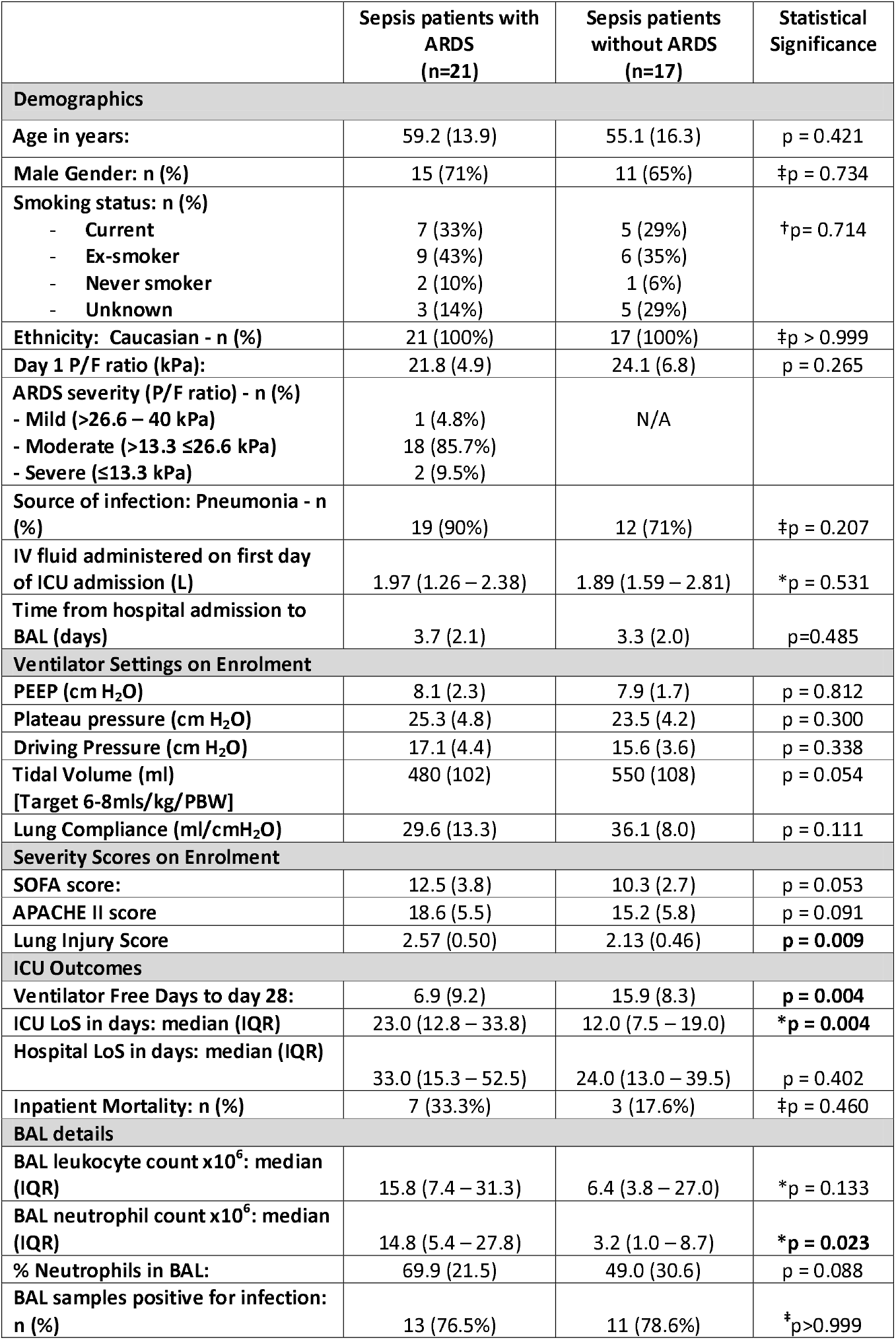
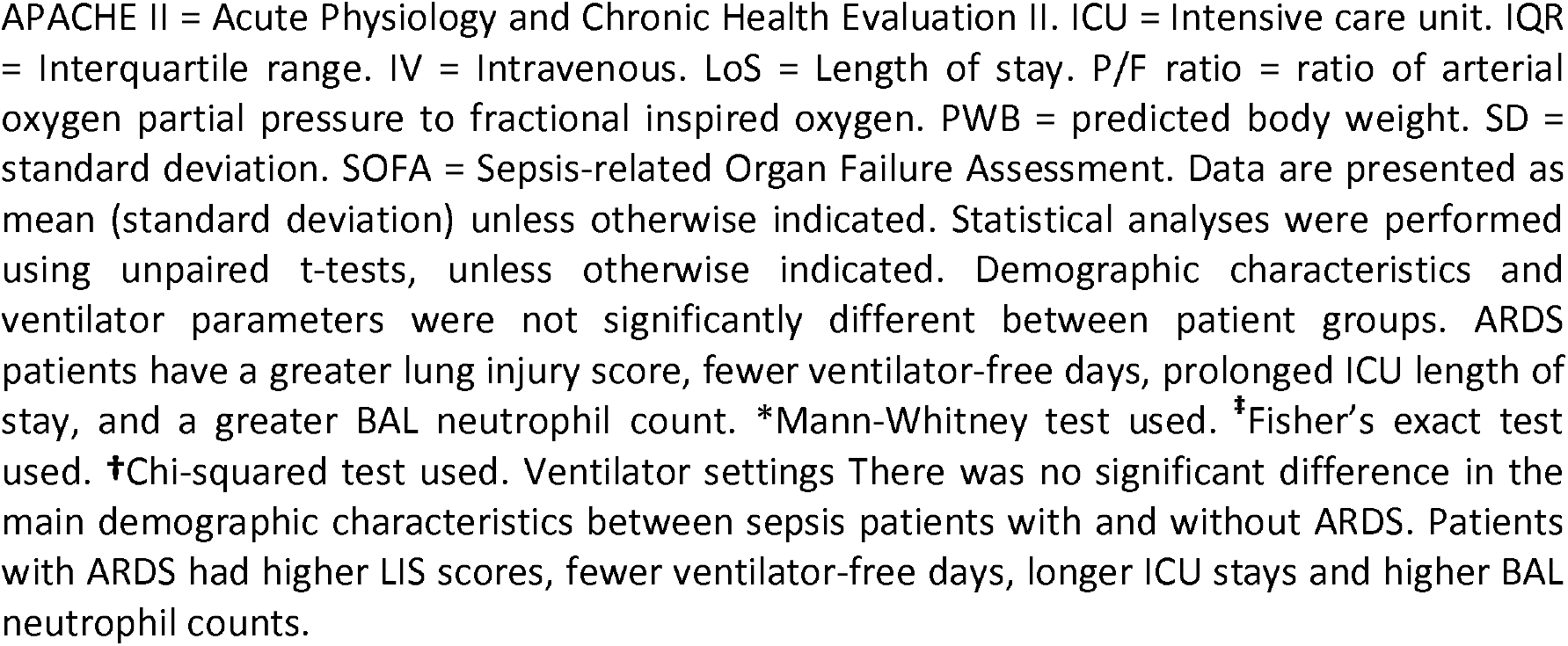
Patient baseline demographic and physiological characteristics.

|  | Sepsis patients with ARDS (n=21) | Sepsis patients without ARDS (n=17) | Statistical Significance |
| --- | --- | --- | --- |
| **Demographics** |  |  |  |
| Age in years: | 59.2 (13.9) | 55.1 (16.3) | p = 0.421 |
| Male Gender: n (%) | 15 (71%) | 11 (65%) | ‡p = 0.734 |
| Smoking status: n (%) |  |  | †p = 0.714 |
| - Current | 7 (33%) | 5 (29%) |  |
| - Ex-smoker | 9 (43%) | 6 (35%) |  |
| - Never smoker | 2 (10%) | 1 (6%) |  |
| - Unknown | 3 (14%) | 5 (29%) |  |
| Ethnicity: Caucasian - n (%) | 21 (100%) | 17 (100%) | ‡p > 0.999 |
| Day 1 P/F ratio (kPa): | 21.8 (4.9) | 24.1 (6.8) | p = 0.265 |
| ARDS severity (P/F ratio) - n (%) |  | N/A |  |
| - Mild (>26.6 – 40 kPa) | 1 (4.8%) |  |  |
| - Moderate (>13.3 ≤26.6 kPa) | 18 (85.7%) |  |  |
| - Severe (≤13.3 kPa) | 2 (9.5%) |  |  |
| Source of infection: Pneumonia - n (%) | 19 (90%) | 12 (71%) | ‡p = 0.207 |
| IV fluid administered on first day of ICU admission (L) | 1.97 (1.26 – 2.38) | 1.89 (1.59 – 2.81) | *p = 0.531 |
| Time from hospital admission to BAL (days) | 3.7 (2.1) | 3.3 (2.0) | p=0.485 |
| **Ventilator Settings on Enrolment** |  |  |  |
| PEEP (cm H<sub>2</sub>O) | 8.1 (2.3) | 7.9 (1.7) | p = 0.812 |
| Plateau pressure (cm H<sub>2</sub>O) | 25.3 (4.8) | 23.5 (4.2) | p = 0.300 |
| Driving Pressure (cm H<sub>2</sub>O) | 17.1 (4.4) | 15.6 (3.6) | p = 0.338 |
| Tidal Volume (ml) | 480 (102) | 550 (108) | p = 0.054 |
| [Target 6-8mls/kg/PBW] |  |  |  |
| Lung Compliance (ml/cmH<sub>2</sub>O) | 29.6 (13.3) | 36.1 (8.0) | p = 0.111 |
| **Severity Scores on Enrolment** |  |  |  |
| SOFA score: | 12.5 (3.8) | 10.3 (2.7) | p = 0.053 |
| APACHE II score | 18.6 (5.5) | 15.2 (5.8) | p = 0.091 |
| Lung Injury Score | 2.57 (0.50) | 2.13 (0.46) | **p = 0.009** |
| **ICU Outcomes** |  |  |  |
| Ventilator Free Days to day 28: | 6.9 (9.2) | 15.9 (8.3) | **p = 0.004** |
| ICU LoS in days: median (IQR) | 23.0 (12.8 – 33.8) | 12.0 (7.5 – 19.0) | **\*p = 0.004** |
| Hospital LoS in days: median (IQR) | 33.0 (15.3 – 52.5) | 24.0 (13.0 – 39.5) | p = 0.402 |
| Inpatient Mortality: n (%) | 7 (33.3%) | 3 (17.6%) | ‡p = 0.460 |
| **BAL details** |  |  |  |
| BAL leukocyte count x10<sup>6</sup>: median (IQR) | 15.8 (7.4 – 31.3) | 6.4 (3.8 – 27.0) | *p = 0.133 |
| BAL neutrophil count x10<sup>6</sup>: median (IQR) | 14.8 (5.4 – 27.8) | 3.2 (1.0 – 8.7) | **\*p = 0.023** |
| % Neutrophils in BAL: | 69.9 (21.5) | 49.0 (30.6) | p = 0.088 |
| BAL samples positive for infection: n (%) | 13 (76.5%) | 11 (78.6%) | ‡p>0.999 |

APACHE II = Acute Physiology and Chronic Health Evaluation II. ICU = Intensive care unit. IQR = Interquartile range. IV = Intravenous. LoS = Length of stay. P/F ratio = ratio of arterial oxygen partial pressure to fractional inspired oxygen. PWB = predicted body weight. SD = standard deviation. SOFA = Sepsis-related Organ Failure Assessment. Data are presented as mean (standard deviation) unless otherwise indicated. Statistical analyses were performed using unpaired t-tests, unless otherwise indicated. Demographic characteristics and ventilator parameters were not significantly different between patient groups. ARDS patients have a greater lung injury score, fewer ventilator-free days, prolonged ICU length of stay, and a greater BAL neutrophil count. \*Mann-Whitney test used. <sup>‡</sup>Fisher's exact test used. <sup>†</sup>Chi-squared test used. Ventilator settings There was no significant difference in the main demographic characteristics between sepsis patients with and without ARDS. Patients with ARDS had higher LIS scores, fewer ventilator-free days, longer ICU stays and higher BAL neutrophil counts.

### Sepsis patients with ARDS have impaired alveolar macrophage efferocytosis and elevated alveolar neutrophil apoptosis

AM efferocytosis was impaired in sepsis patients with ARDS compared to those without ARDS (Figure 1A, means 7.6 vs 22.7%, p=0.002) and lobectomy patients (Figure 1A, means 7.6 vs 32.2%, p<0.0001). There was no difference between AM efferocytosis in sepsis patients without ARDS and lobectomy patients (Figure 1A). AM phagocytosis was also assessed, however n-numbers were limited due to modest BAL AM yields. There was no difference in AM phagocytosis of *E. coli* and *S. Aureus* pHrodo® bio-particles between sepsis patients with and without ARDS (supplemental figure 3). Alveolar neutrophil apoptosis (assessed immediately post-BAL) was greater in sepsis patients with ARDS compared to those without ARDS (Figure 1B, means 41.3% vs 14.1%, p=0.0001). A trend towards increased alveolar neutrophil necrosis in sepsis patients with ARDS compared to those without ARDS was observed, but this did not reach statistical significance (4.5% vs 1.1%, p=0.191).

**Figure 1:**
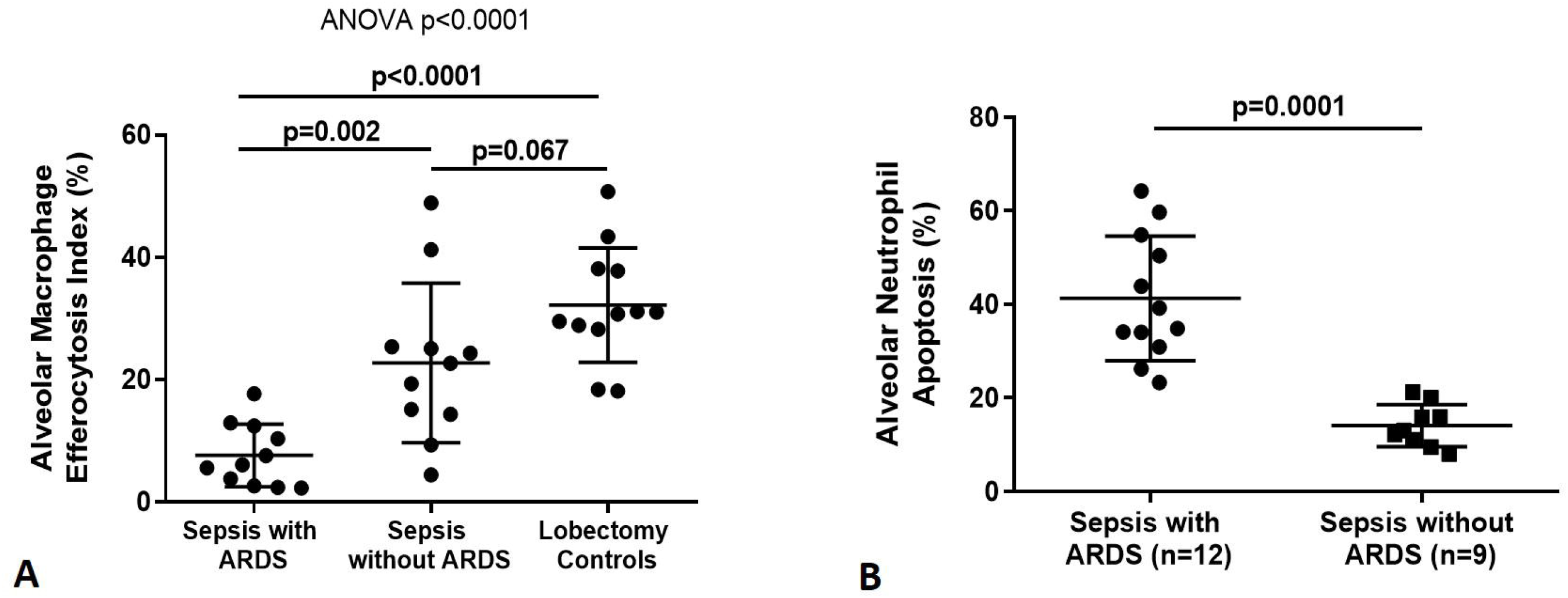
Alveolar macrophage efferocytosis and BAL neutrophil apoptosis in sepsis patients with and without ARDS. **A:** Alveolar macrophages (AMs) from sepsis patients with ARDS have significantly reduced efferocytosis index compared to sepsis patients without ARDS (means 7.6 vs 22.7%, p=0.002) and lobectomy controls (means 7.6 vs 32.2%, p<0.0001). Statistical analysis by ANOVA and Dunns multiple comparisons test, n=11-12 in all groups. **B:** Neutrophil apoptosis assessed within 1 hour of broncho-alveolar lavage (BAL) fluid collection. Sepsis patients with ARDS have significantly greater percentage of apoptotic neutrophils in BAL compared to sepsis patients without ARDS (medians 41.3 vs 14.1%, p=0.0001). Error bars shown as mean and standard deviation. Statistical analysis by Welch’s t-test. Some patients did not have neutrophil apoptosis measured since this was only added to the study protocol after recruitment had already begun.

### Impaired alveolar macrophage efferocytosis is associated with increased alveolar inflammation and worse clinical outcomes in ventilated sepsis patients

Across all sepsis patients (with and without ARDS), AM efferocytosis showed negative correlation with BAL concentrations of IL-8 (Figure 2A, p=0.0003) and IL-1ra (Figure 2B, p=0.004). There was no correlation between AM efferocytosis and BAL concentrations of IL-6, IL-1β, TNF-α, IL-10, VEGF and MCP-1 (data not shown, p>0.05 following Bonferroni correction).

**Figure 2:**
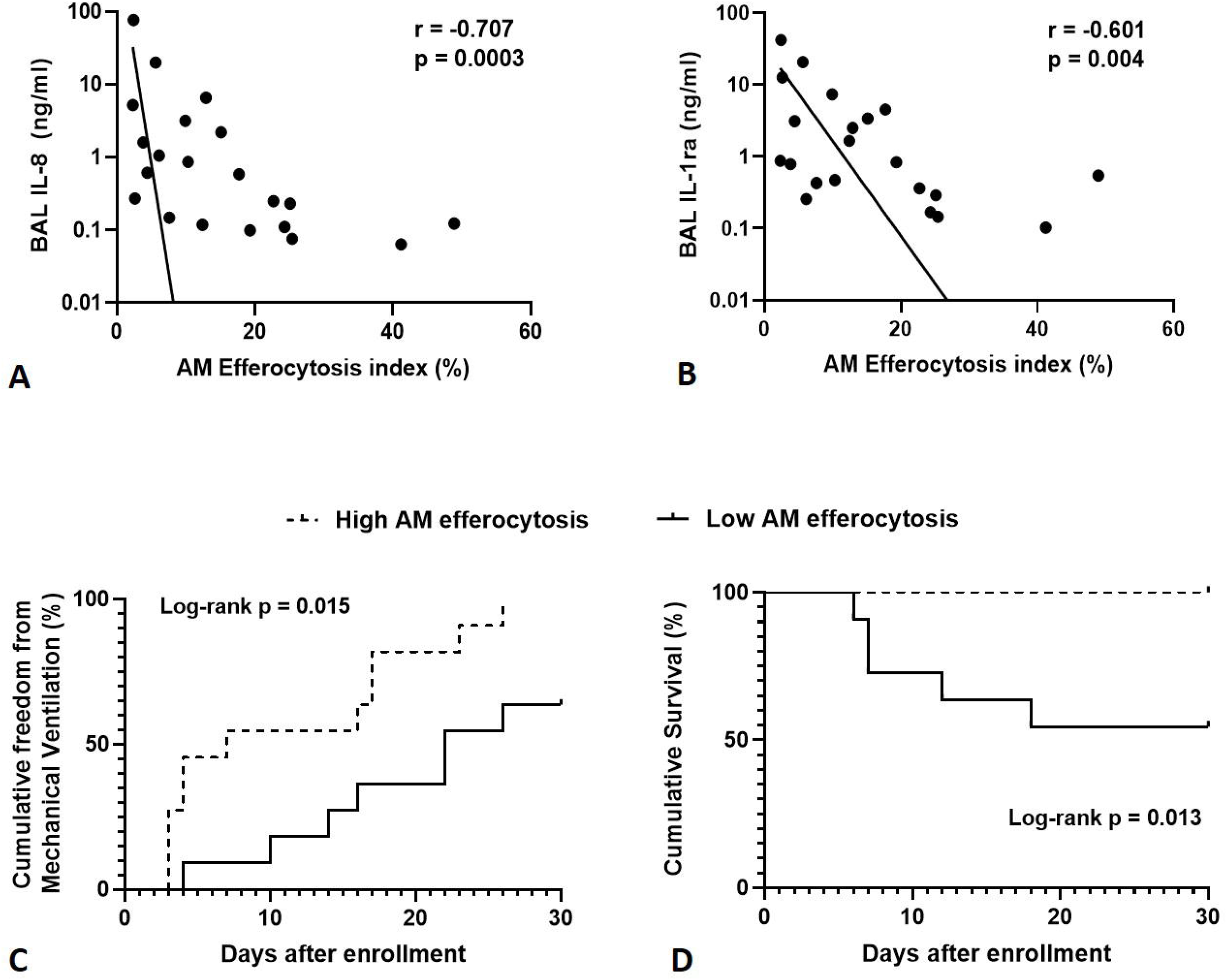
Correlations between alveolar macrophage efferocytosis, alveolar cytokines and clinical outcomes in ventilated sepsis patients. Levels of IL-1ra and IL-8 were measured in BAL from sepsis patients with and without ARDS, then correlated with AM efferocytosis index. **A-B:** There is significant negative correlation between alveolar macrophage efferocytosis index and BAL concentrations of IL-8 (r = -0.707, p=0.0003) and IL-1ra (r = -0.601, p=0.004) in sepsis patients with and without ARDS. BAL = broncho-alveolar lavage fluid. IL-1ra = Interleukin 1 receptor antagonist. Log scales used for both graphs, semi-log non-linear line of fit used, n=21 for both plots. Spearman’s correlation coefficient with Bonferroni’s correction used for statistical analysis. Bonferroni corrected significance p<0.00625. **C:** A threshold AM efferocytosis index of 12.7% was used to distinguish between ‘low’ and ‘high’ efferocytosis, based on this value being 1 standard deviation above the mean AM efferocytosis index of sepsis patients with ARDS. For all sepsis patients (with and without ARDS), low AM efferocytosis index was associated with reduced freedom from mechanical ventilation in the 30 days from enrolment (χ^2^ = 7.41, p=0.015). Statistical analysis by log-rank test, n=11 in each group. **D:** For all sepsis patients (with and without ARDS), low AM efferocytosis index was associated with decreased survival in the 30 days from enrolment (χ ^2^ = 6.22, p=0.013). Statistical analysis by log-rank test, n=11 in each group.

For all sepsis patients (with and without ARDS), low AM efferocytosis index was associated with reduced freedom from mechanical ventilation in the 30 days from enrolment (Figure 2C, p=0.015). For all sepsis patients (with and without ARDS), low AM efferocytosis index was associated with increased mortality in the 30 days from enrolment (Figure 2D, p=0.013).

### ARDS BAL treatment of alveolar macrophages impairs efferocytosis and preserves bacterial phagocytosis

Due to modest BAL AM yields in ARDS patients, the necessary experiments could not all be performed on each enrolled patient. We had identified an association between impaired AM efferocytosis and intense alveolar inflammation (Figure 1A-B). Therefore, we sought to utilize an *in vitro* model of ARDS, by treating lobectomy patient AMs with pooled ARDS BAL to induce AM dysfunction. BAL samples from the first 14 ARDS patients recruited were pooled and characterized with regards to inflammatory cytokine and lipopolysaccharide content (supplemental table 2). ARDS BAL or saline vehicle control (VC) treatment had no effect on AM apoptosis or viability (supplemental figure 4).

ARDS BAL treatment of AMs reduced efferocytosis compared to VC treatment (Figure 3A, p=0.0006). ARDS BAL treatment of AMs reduced Rac1 mRNA expression compared to VC treatment (Figure 3B, p=0.016), which supports our finding that ARDS BAL inhibits AM efferocytosis.

**Figure 3:**
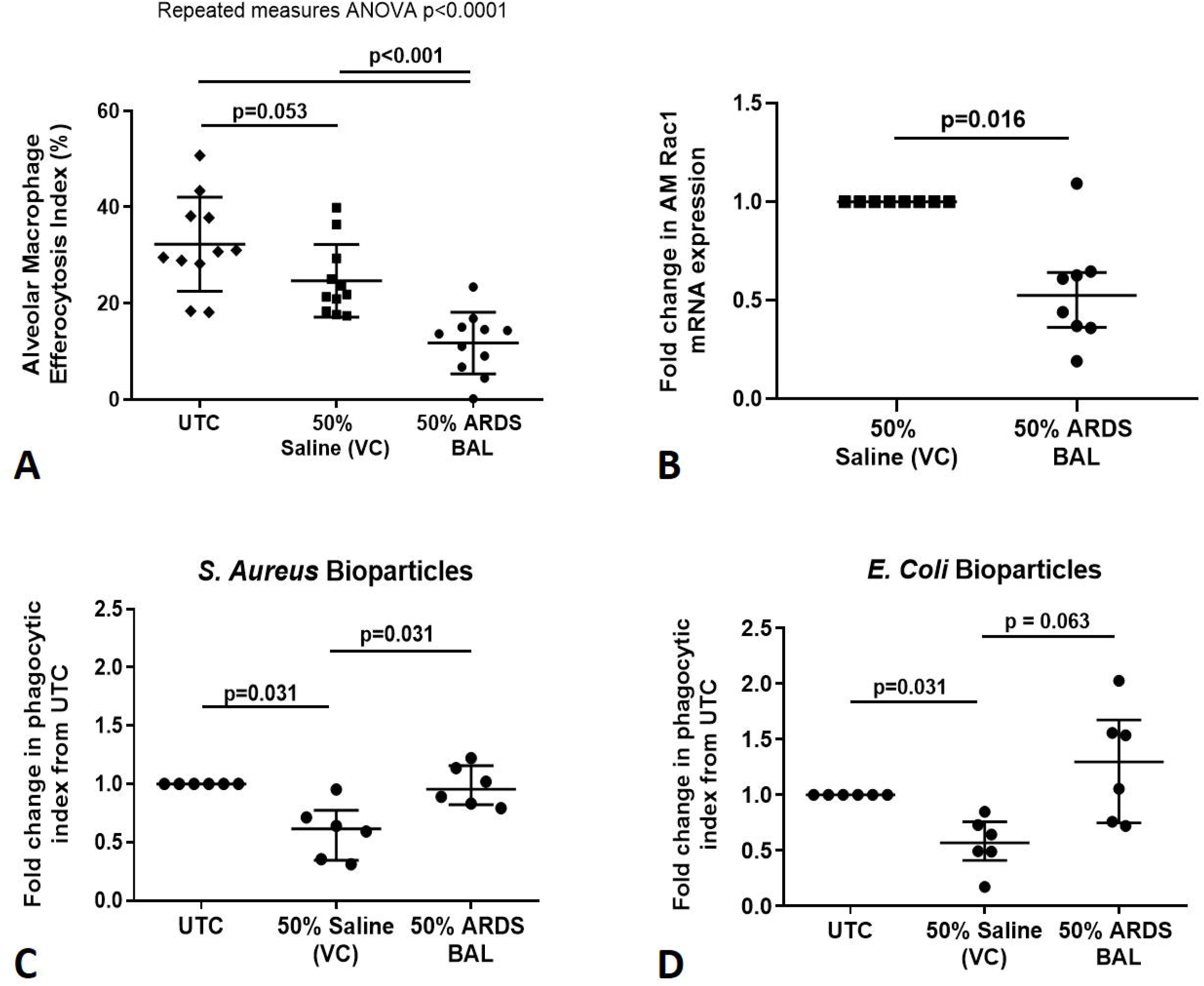
Effect of pooled ARDS BAL treatment on lobectomy patient alveolar macrophage efferocytosis and phagocytosis. **A:** UTC = Untreated Control (cultured in RPMI + 10% FBS). VC = Vehicle control (50% saline). Effect of ARDS BAL treatment on lobectomy patient AM efferocytosis. Treatment with 50% ARDS BAL significantly reduced lobectomy patient AM efferocytosis compared to VC treatment (mean of differences 13.0%, p=0.0006) and UTC (mean of differences 20.6%, p=0.0009). Treatment of AMs with VC had no effect on efferocytosis compared to UTC (mean of differences 7.6%, p=0.053). Statistical analysis by repeated measures ANOVA and Tukey’s multiple comparisons test, n=11 for all groups. **B:** Effect of ARDS BAL treatment on Rac1 gene transcription in lobectomy patient AMs. Data shown as fold change in AM Rac1 mRNA expression from 50% saline treatment. Statistical analysis by Wilcoxon matched-pairs signed rank test, n=8. VC = vehicle control (50% saline). Treatment with 50% ARDS BAL significantly reduced Rac1 mRNA expression in lobectomy patient AMs, compared to VC treatment (median of differences 0.48, p=0.016). Error bars shown as mean and standard deviation. **C-D:** AM phagocytic index in lobectomy AMs treated with 50% ARDS BAL. Data corrected to fold change in phagocytic index from untreated control (UTC – cultured in RPMI + 10% FBS). Statistical analysis by Wilcoxon matched-pairs signed rank test, n=6 for all groups, *p<0.05. VC = Vehicle control (50% Saline). VC values are non-identical in graphs C and D. Data shown as median and interquartile range. **C:** Treatment of AMs with 50% saline vehicle control reduced phagocytosis of *S. Aureus* bioparticles (median of differences -0.38, p = 0.031). Treatment with 50% ARDS BAL caused a significant increase in AM phagocytosis of *S. Aureus* bioparticles compared to VC (median of differences 0.32, p=0.031). **D:** Treatment of AMs with 50% saline vehicle control reduced phagocytosis of *E. coli* bioparticles (median of differences -0.43, p = 0.031). No significant difference in in AM phagocytosis of *E. coli* bioparticles was observed following treatment with 50% ARDS BAL compared to VC (median of differences 0.59, p=0.063).

ARDS BAL treatment of AMs increased phagocytosis of *S. Aureus* pHrodo® bioparticles compared to VC treatment (Figure 3C, p=0.031). There was no difference in AM phagocytosis of *E. coli* pHrodo® bioparticles following ARDS BAL treatment compared to VC treatment (Figure 3D, p=0.063). VC treatment of AMs reduced phagocytosis of both *S. Aureus* and *E. coli* pHrodo® bioparticles compared to untreated controls (Figure 3C-D, p=0.031). Thus, ARDS BAL treatment had divergent effects on AM efferocytosis and phagocytosis.

### ARDS BAL treatment of alveolar macrophages alters surface receptor expression

Since ARDS BAL treatment of AMs had divergent effects on efferocytosis and phagocytosis, we used this *in vitro* model to investigate the association between AM phenotype and function. ARDS BAL treatment of lobectomy patient AMs increased expression of CD206 (Figure 4A, p=0.006) and MerTK (Figure 4B, p=0.028) compared to VC treatment. ARDS BAL treatment of AMs decreased SIRPα expression compared to VC treatment (Figure 4C, p=0.006). ARDS BAL treatment of AMs did not change expression of CD163 or CD80 compared to VC treatment (Figure 4D-E, p>0.05 for both). These changes in AM surface receptor expression were incongruent with the observed defect in AM efferocytosis following ARDS BAL treatment. In contrast, treatment of lobectomy patient AMs with a pro-inflammatory mixture of interferon-γ and lipopolysaccharide impaired efferocytosis; however expression of SIRPα was increased, and expression of both MerTK and CD163 was decreased (supplemental figure 5). This indicates that the effect of ARDS BAL treatment on AMs can not solely be explained by changes in surface receptor expression.

**Figure 4:**
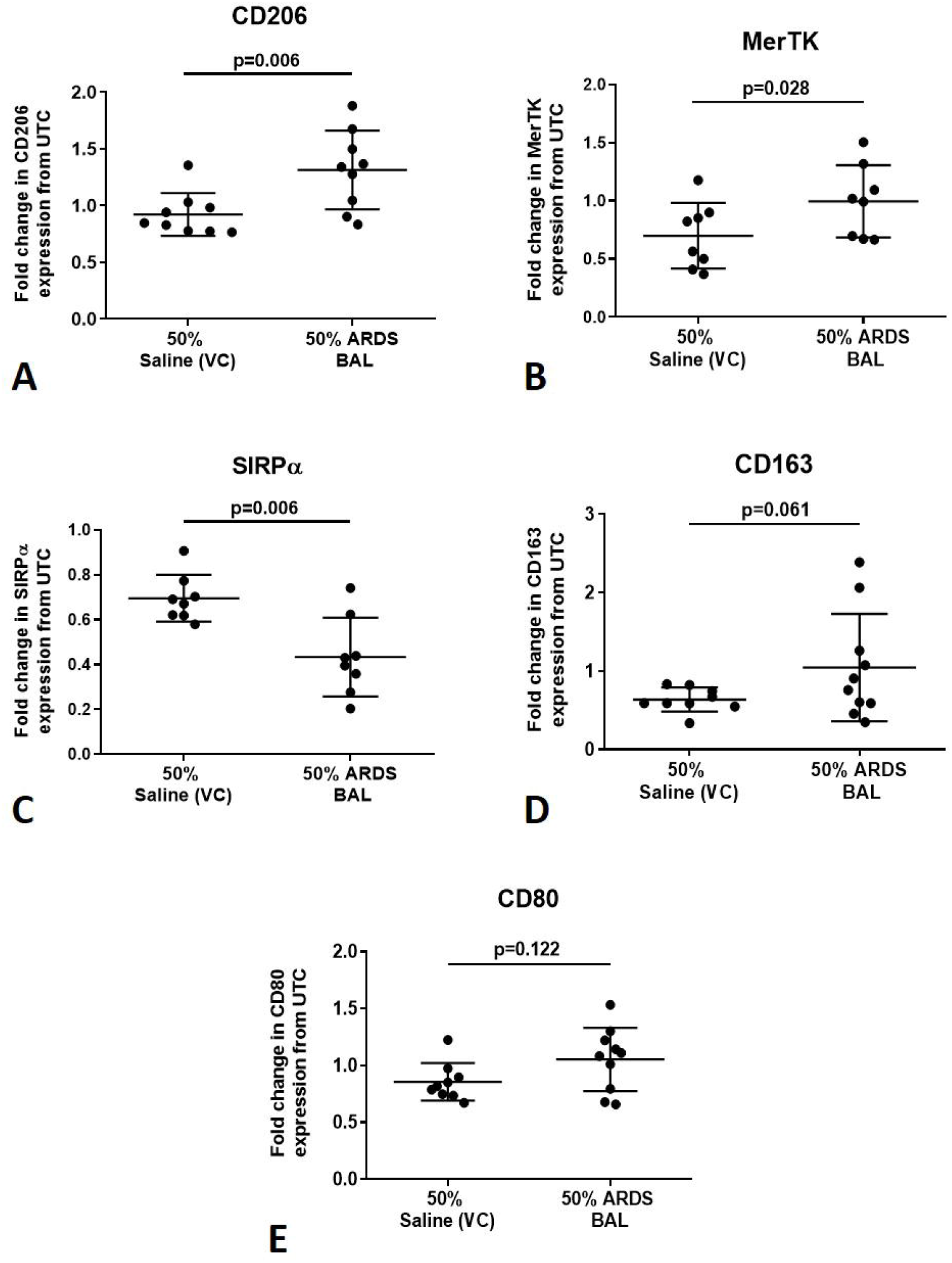
Effect of ARDS BAL treatment on lobectomy patient alveolar macrophage surface receptor expression. UTC = Untreated control (RPMI + 10% FBS). VC = Vehicle control (50% Saline). Mer = Mer receptor tyrosine kinase. SIRPα = Signal regulatory protein alpha. Statistical analysis by paired t-test, n≥8 for all groups. **A:** 50% ARDS BAL treatment significantly increased CD206 expression on AMs compared to VC treatment (mean of differences 0.39, p=0.006). **B:** 50% ARDS BAL treatment significantly increased Mer expression on AMs compared to VC treatment (mean of differences 0.30, p=0.028). **C:** 50% ARDS BAL treatment significantly decreased SIRPα expression on AMs compared to VC treatment (mean of differences 0.26, p=0.006). **D:** 50% ARDS BAL treatment increased CD163 expression on AMs compared to VC treatment, however this difference did not reach statistical significance (mean of differences 0.44, p=0.061). E: 50% ARDS BAL treatment did not significantly change CD80 expression on AMs compared to VC treatment (mean of differences 0.19, p=0.122).

### ROCK-inhibition partially restores alveolar macrophage efferocytosis in an in vitro ARDS model

Since ARDS BAL treatment downregulated Rac1 gene expression in AMs, we sought to upregulate Rac1 expression and restore efferocytosis by inhibiting ROCK and PTEN. Rac1 intracellular signalling pathways are summarised in Figure 5. Addition of ROCK-inhibitor to ARDS BAL treatment increased AM efferocytosis compared to treatment with ARDS BAL plus VC (Figure 6A, p=0.009). Addition of PTEN-inhibitor to ARDS BAL treatment had no effect on efferocytosis compared to treatment with ARDS BAL plus VC (Figure 6B). ROCK inhibition had no effect of AM phagocytosis or phenotype (supplemental figures 6-7).

**Figure 5:**
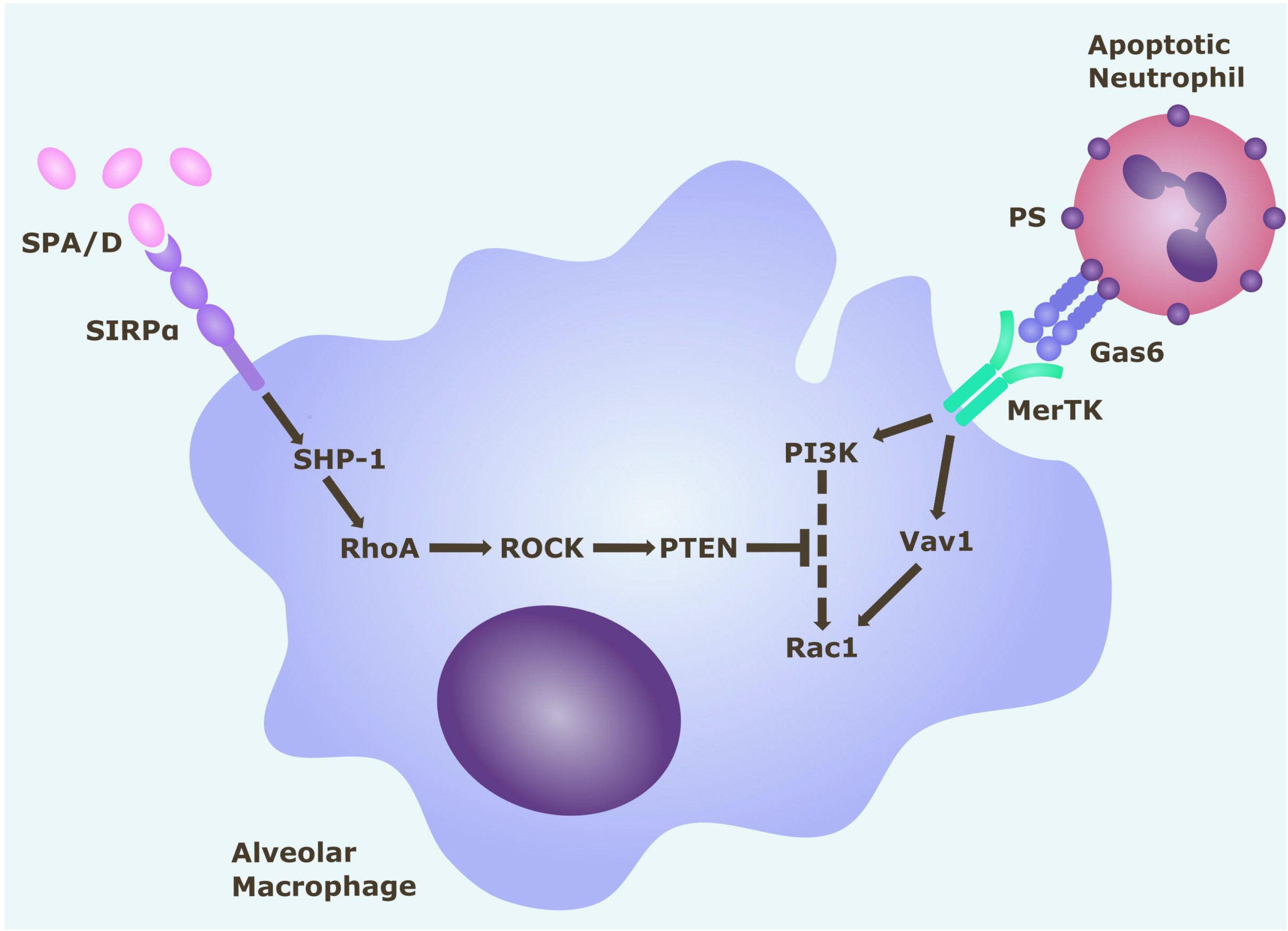
Rac1 intracellular signalling pathways in alveolar macrophages. Alveolar macrophage efferocytosis is regulated by surface receptors MerTK and SIRPα. Gas6 binds to PS on the surface of apoptotic cells. Activation of MerTK by the PS opsonin Gas6 can trigger signalling cascades via PI3K and Vav1, which both upregulate Rac1. Activation of Rac1 results in cytoskeletal re-arrangement and efferocytosis of the apoptotic cell. Activation of SIRPα by SP-A (or SP-D) triggers a signalling cascade along the SHP1/RhoA/ROCK/PTEN pathway, which inhibits PI3K signalling, and ultimately downregulates Rac1, thereby inhibiting efferocytosis. Gas6 = growth arrest specific-6. MerTK = Mer tyrosine kinase receptor. PI3K = phosphatidylinositol 3’-OH kinase. PS = phosphatidylserine. PTEN = phosphatase and tensin homologue. ROCK = Rho-associated kinase. SHP-1 = Src homology region 2 domain-containing phosphatase-1. SIRPα = signal regulatory protein-α. SPA/D = surfactant protein A / D.

**Figure 6:**
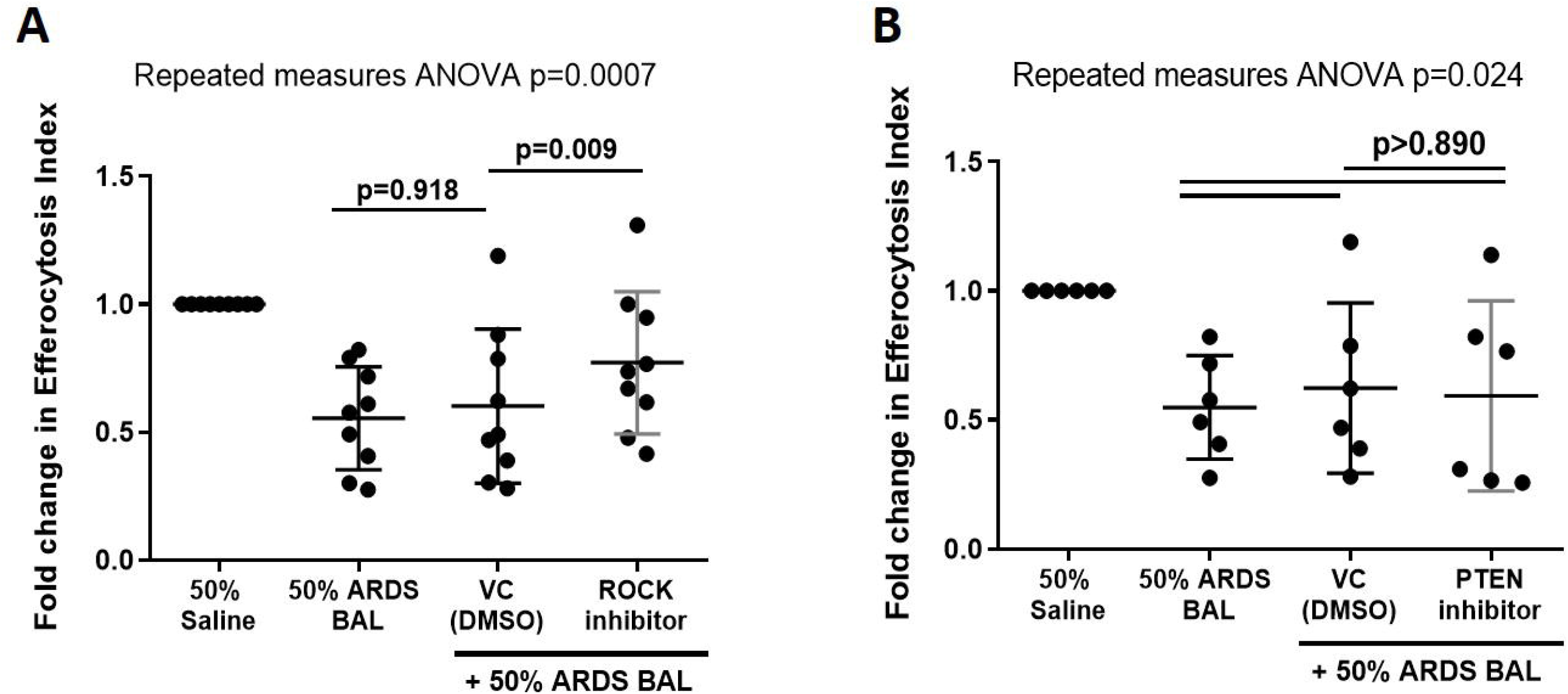
Effect of ROCK and PTEN inhibitors on restoring alveolar macrophage efferocytosis following ARDS BAL treatment. Data shown as fold change in alveolar macrophage (AM) efferocytosis index from 50% saline treatment. ROCK = Rho-associated protein kinase. PTEN = Phosphatase and tensin homolog. VC = Vehicle Control (Dimethyl Sulfoxide [DMSO] at 1:50,000 dilution). ROCK-inhibitor = 200nM Y-27632 dihydrochloride. PTEN-inhibitor = 2μM SF1670. Statistical analysis by repeated measures ANOVA with Tukey’s multiple comparison test. **A:** Addition of VC to ARDS BAL mixture had no significant effect on AM efferocytosis (mean of differences 0.05, p=0.918, n=9) compared to ARDS BAL alone. Addition of ROCK-inhibitor to ARDS BAL treatment significantly increased efferocytosis compared to treatment with VC + ARDS BAL (mean of differences 0.17, p=0.009, n=9) **B:** Addition of PTEN-inhibitor to 50% ARDS BAL treatment had no significant effect on efferocytosis compared to treatment with VC + ARDS BAL (mean of differences 0.03, p=0.924, n=6).

## DISCUSSION

The main findings from our study can be summarized as follows: 1) Sepsis patients with ARDS have impaired AM efferocytosis and elevated alveolar neutrophil apoptosis, compared to control ventilated sepsis patients. 2) Across all ventilated sepsis patients (with and without ARDS), impaired AM efferocytosis is associated with intense alveolar inflammation, prolonged invasive ventilation and mortality. 3) ARDS BAL treatment of lobectomy patient AMs induced an impairment in efferocytosis similar to that observed in ARDS patients, but preserved AM bacterial phagocytosis. Thus, the inflammatory contents of ARDS BAL do not induce a global impairment in AM function, but rather a specific impairment in efferocytosis. 4) The impairment in AM efferocytosis caused by ARDS BAL treatment was mediated by suppression of intracellular Rac1 expression, and not by changes in surface receptor expression. 5) ROCK-inhibition partially restored AM efferocytosis in the *in vitro* model of ARDS. Modulation of the ROCK-PI3K-Rac1 intracellular signalling pathway may offer a therapeutic strategy to upregulate AM efferocytosis in ARDS.

A previous study published in this journal(17) showed that circulating monocytes from ARDS patients, differentiated to macrophages *ex vivo*, had impaired efferocytosis. Our study identified an impairment in AM efferocytosis, which supports these findings and is more relevant to the pathogenesis of ARDS, since the disease process originates in the alveolar space. The control group in our study consisted of ventilated sepsis patients, in contrast to this previous study, which included control patients healthy enough to undergo outpatient bronchoscopy(17). Therefore, our study was able to determine that the decrease in AM efferocytosis associated with the development of sepsis-related ARDS, occurs independently of sepsis, ICU admission and invasive ventilation.

In early ARDS, pro-inflammatory monocytes migrate to the alveoli, then differentiate into ‘recruited’ AMs(22). A direct correlation was observed between alveolar monocyte influx, severity of respiratory failure and mortality(22). Murine models showed that following initiation of lung injury, the majority of inflammatory cytokines were released by recruited AMs(23). Our study assessed global AM efferocytosis, and did not distinguish between resident and recruited AMs. Therefore, we initially postulated that the decreased AM efferocytosis in ARDS may be due to polarization of AMs to a pro-inflammatory phenotype, which is associated with reduced efferocytosis(24). We then undertook experiments using the *in vitro* model of ARDS to investigate the association between AM phenotype and function.

Intriguingly, ARDS BAL treatment of AMs increased expression of efferocytosis receptors (CD206 and MerTK), and decreased expression of the anti-efferocytosis receptor SIRPα. These phenotypic changes were incongruent with the functional defect in AM efferocytosis induced by ARDS BAL. In comparison, treatment with pro-inflammatory mediators (IFNγ and lipopolysaccharide) also decreased AM efferocytosis, but induced SIRPα expression and decreased MerTK expression. Although ARDS BAL and pro-inflammatory mediator treatments both impaired AM efferocytosis, they had opposite effects on efferocytosis receptor expression. A similar association was observed in cigarette smokers, between decreased AM efferocytosis(25), increased MerTK expression(26), and increased transcription of genes associated with alternative activation(27). COPD patients also have impaired AM efferocytosis(28) with overexpression of efferocytosis receptors CD206 and CD163 (29). Our data therefore suggest that the AM efferocytosis defect induced by ARDS BAL treatment is not mediated by receptor changes, but by changes in the expression of intracellular signalling mediators such as Rac1. Further analysis of ARDS BAL is required to identify other components (e.g. microRNA transfer via extracellular vesicles) which may affect the intracellular pathways regulating AM efferocytosis(30).

Strategies to upregulate AM efferocytosis may reduce secondary necrosis of alveolar neutrophils, thereby attenuating inflammation in ARDS. Since *in vitro* ARDS BAL treatment downregulated AM Rac1 gene expression, we sought to upregulate Rac1 expression and restore efferocytosis by inhibiting ROCK and PTEN. Addition of ROCK-inhibitor to ARDS BAL treatment partially restored AM efferocytosis function, and had no effect on bacterial phagocytosis. This supports our theory that modulation of the ROCK-PI3K-Rac1 intracellular signalling pathway is a mechanism by which ARDS BAL suppresses AM efferocytosis. However addition of PTEN-inhibitor had no significant effect; this may be because the role of PTEN is less important in antagonizing the PI3K pathway. ROCK-inhibitors have been shown to increase efferocytosis in MDMs and AMs from COPD patients(31). Further studies to investigate the role of the ROCK-PTEN-Rac1 pathway in ARDS AM dysfunction are required. ROCK inhibition promotes PI3K signalling, which has multiple effects on cellular function beyond upregulation of Rac1, including proliferation, chemotaxis, and migration(32). ROCK inhibition would have many off-target effects, thereby limiting its therapeutic potential as a strategy to upregulate AM function in ARDS. Existing medications could be tested using the *in vitro* model of ARDS, to determine if they can restore AM efferocytosis e.g. N-acetylcysteine(33), macrolide antibiotics(34), statins(35) and glucocorticoids(36).

The accumulation of apoptotic alveolar neutrophils we observed in ARDS patients could be due to increased apoptosis and / or decreased clearance. In contrast, previous studies have shown that ARDS BAL treatment of neutrophils from healthy volunteers delays apoptosis following *ex-vivo* culture for 18-24 hours(17, 37-39). Alveolar and circulating neutrophils from ARDS patients also showed delayed apoptosis following *ex vivo* culture for 20 hours, compared to circulating neutrophils from healthy volunteers(39). However, these results cannot be directly compared with those from our study, in which alveolar neutrophil apoptosis was assessed immediately after BAL (without *ex vivo* culture). Only two studies have previously investigated alveolar neutrophil apoptosis in early ARDS immediately after BAL (37, 38). Neither study showed a significant difference in alveolar neutrophil apoptosis between ARDS and control at-risk patient groups; furthermore the trends observed were contradictory. Therefore, only limited comparisons can be drawn with results from our study. Our data suggest that alveolar neutrophil apoptosis is initially delayed due to the pro-inflammatory contents of ARDS BAL, however once apoptosis occurs the neutrophils persist due to impaired AM efferocytosis.

Our study had some limitations. ARDS patient recruitment was limited because prior steroid use, immunosuppression (e.g. post-transplant) or immunodeficiency were all exclusion factors. Even if meeting the inclusion criteria, some patients could not safely undergo bronchoscopy, due to their ventilation status; no patients with severe ARDS were recruited for this reason. Therefore, patient recruitment numbers were modest. Following bronchoscopy on ARDS patients, the BAL was often highly neutrophilic, making it difficult to isolate AMs. The average number of BAL AMs isolated was 1.2 million, meaning that often only one functional assay could be performed per patient; efferocytosis assays were given priority. Due to logistical constraints, efferocytosis assays were undertaken with heterologous neutrophils, as opposed to autologous neutrophils, which would have more accurately reflected the environment *in vivo*. Owing to difficulties associated with isolating AMs from ARDS patients, the *in vitro* model of ARDS was set up using AMs from lung resection tissue and pooled ARDS BAL to recapitulate the AM dysfunction observed *in vivo*. Although unaffected lung tissue was processed, we cannot rule out contamination with tumour-associated macrophages which are characterised by an immunosuppressive phenotype, and may exhibit increased efferocytosis(40), which could account for some of the divergent effects observed. Two different sources of AMs were utilised, with lobectomy AMs obtained after most BAL AMs, which could have introduced a small bias.

In summary, our findings indicate that patients with sepsis-related ARDS have impaired AM efferocytosis, which potentially contributes to ARDS pathogenesis and negatively impacts clinical outcomes, including mortality. Strategies to upregulate AM efferocytosis may be of value for attenuating inflammation in ARDS.

## Data Availability

Anonymised data are available upon reasonable request to the corresponding author.

## Acknowledgements

We would like to thank the nurses and thoracic surgeons of the Thoracic Research Team, University Hospitals Birmingham NHS Foundation Trust for their help consenting patients undergoing thoracic surgery. We also thank Dr Gerald Langman and Dr Andrew Robinson, Department of Cellular Pathology, University Hospitals Birmingham NHS Foundation Trust for their help in obtaining lung tissue samples from patients undergoing lobectomy.

## Contributors

RYM, AS, GGL, MAM, GDP, and DRT contributed to study conception and design. RYM, AS, DP, STL, and RSH contributed to data acquisition. All authors contributed to the data analysis and interpretation. RYM, AS, GDP and DRT drafted the manuscript. All authors critically revised the manuscript for intellectual content and approved the final version before submission. RYM and AS share joint first authorship. GDP and DRT share joint senior authorship.

## Data Availability Statement

Anonymised data are available upon reasonable request to the corresponding author.

