## Supplemental for "Acute Respiratory Distress Syndrome is associated with impaired alveolar macrophage efferocytosis"

**Online Supplement**

**Methods (supplemental)**

Patient severity scores

The Sepsis-related Organ Failure Assessment (SOFA) Score was used to determine the severity of organ failure suffered by critically ill patients with a septic aetiology (1, 2). Organ function in respiratory, neurological, cardiovascular, haematological, hepatic and renal systems are given a score between 0 - 4. Greater total scores are associated with worse clinical outcomes.

The Acute Physiology and Chronic Health Evaluation II (APACHE II) system was used as a marker of disease severity in critically ill patients. In APACHE II, 12 physiologic variables are each assigned a score from 0 - 4 depending on the most abnormal measurement in the first 24 hours of their ICU stay. Additional scores are given for variables including age and concurrent medical or surgical conditions. This results in a score between 0 - 71, with greater scores indicating greater disease severity and consequently increased risk of mortality (3).

The Murray Lung Injury Score (LIS) was developed prior to the Berlin and Consensus criteria for ARDS diagnosis, in order to aid identification of acute lung injury(4). LIS components include alveolar infiltrates on chest x-ray, PEEP, PaO_2_/FiO_2_ ratio and lung compliance.

Alveolar Macrophage Isolation

Non-affected, macroscopically normal lung tissue samples were perfused with 0.15M saline via pressure bag by inserting a needle (21-gauge) in bronchioles. When saturated, the tissue was gently massaged to facilitate emptying lavage fluid from the tissue, ready for the next instillation. This process was repeated until the lavage fluid contained fewer than 1x10^4^ cells/ml (5). From this stage onwards, patient BAL and lung tissue lavage fluid were processed identically.

Cells were pelleted from the BAL / lavage fluid by centrifugation at 500g for 5 minutes. Mononuclear cells were then separated by gradient centrifugation using Lymphoprep (StemCell Technologies), according to the manufacturer’s instructions. Mononuclear cells were then cultured in RPMI-1640 media supplemented with 10% Foetal Calf Serum (FCS), 100U/mL penicillin, 100ug/mL streptomycin and 2mM L-glutamine (Sigma-Aldrich) at 37^o^C and 5% CO_2_ for 24 hours. During this time, alveolar macrophages adhered to the plastic wells. After 24 hours culture the wells were washed and media changed, thereby removing non-adherent mononuclear cells (6, 7). Alveolar macrophages were then assessed for purity by Cytospin (5). Alveolar macrophage purity was consistently >95% across all samples.

Alveolar Macrophage Efferocytosis Assay

The efferocytosis assay was modified from published protocols (8-11). Neutrophils were isolated from the blood of healthy volunteers using Percoll density centrifugation (12) as previously described by our group (13). Neutrophil purity was >96% as assessed by cytospin, and viability >97% as assessed by trypan blue exclusion. Neutrophils were suspended in a 5μM solution of CellTracker^TM^ Deep Red fluorescent dye (ThermoFisher Scientific) in 10% FCS/RPMI at 4 x 10^6^ /ml, then incubated for 30 minutes at 37^0^C. CellTracker^TM^ Deep Red has an excitation/emission spectra of 630/650 nm maxima; similar to that of allophycocyanin (APC). Stained neutrophils were centrifuged at 1500g for 5 minutes then re-suspended at 2 x 10^6^ / ml in serum-free RPMI and incubated at 37^0^C and 5% CO_2_ for 24 hours to allow apoptosis. Flow cytometric assessment of neutrophil apoptosis was performed using a Fluorescein Isothiocyanate (FITC)-conjugated Annexin V and 7-aminoactinomycin D (7-AAD) apoptosis detection kit (Biolegend): mean neutrophil apoptosis of 93% with necrosis of <2% was observed.

AMs were cultured at 2.5 x10^5^ /well in 24-well plates. As negative control, 5μg/ml Cytochalasin D (CytoD, Sigma-Aldrich) was added for 30 minutes to inhibit actin filament polymerization required for efferocytosis. Stained apoptotic neutrophils (ANs) were added to AMs at a 4:1 ratio prior to incubation for 2 hours at 37^o^C. The optimal assay duration of 2 hours had previously been determined by time course experiments. Media was removed and wells washed twice with ice-cold PBS to remove non-adherent/engulfed neutrophils. Cells were harvested using a 5 minute TrypLE™ express (ThermoFisher) incubation at 37^o^C, prior to acquisition using an Accuri C6 flow cytometer and software (BD Biosciences). AMs and ANs alone were used to set gates for their respective populations on forward and side-scatter plots. ANs alone were used to set a positive gate on the APC plot, which was subsequently used to identify AMs which had engulfed ANs. Minimum 5,000 events gated as AMs were counted for each experimental condition and the percentage of APC^+^ AMs calculated. CytoD treated AMs (negative control) determined the background fluorescence present due to ANs adhering to the surface of AMs, but not being engulfed. This background fluorescence was subtracted from the percentage of APC^+^ AMs in other experimental conditions, to give a corrected net efferocytosis index representative of neutrophil engulfment (supplemental figure 1). Steps were taken to avoid bias, including drawing gates based on single cell populations (ANs and AMs) prior to assessing efferocytosis.

AM Phagocytosis Assay

AM Phagocytosis assays were performed using pHrodo^TM^ red *E. Coli* and *S. Aureus* BioParticle^®^ conjugates (ThermoFisher Scientific) in a 96 well plate according to manufacturer’s instruction and as previously described (5). pHrodo^TM^ beads were prepared according to manufacturer’s instructions at a final concentration of 1 mg/ml. AMs were seeded at 50,000 cells per well in black well, clear bottomed 96 well plates (VWR, Sussex, UK) and allowed to adhere overnight. For negative control wells, 5μg/ml Cytochalasin D (Sigma-Aldrich) was added for 30 minutes to inhibit actin filament polymerization, thus preventing macrophage phagocytosis. 50 μL of pHrodo bead suspension was added per well and incubated for 6 hours at 37°C. After 6 hours, cells were washed three times with PBS before adding of 100 μl fresh PBS for reading. Fluorescence was measured using a microplate reader (Synergy 2, Bio-Tek, USA) set at the excitation / emission spectra of pHrodo^TM^ red dye: 560 / 585 nm. The negative control (cytochalasin D treated) AMs were used to determine the background fluorescence present due to stained pHrodo^TM^ red BioParticles^®^ adhering to the outside of macrophages, but not being engulfed (since cytochalasin D inhibits phagocytosis). This background fluorescence value was subtracted from fluorescence values of other experimental conditions, to give corrected net fluorescence value in representative of phagocytosis.

For AMs derived from patient BAL, phagocytosis was calculated and expressed as relative fluorescence units (RFU). For AMs derived from lung resections and treated with ARDS BAL, phagocytosis results were expressed as fold change in RFU from untreated AMs.

Use of Alveolar Macrophages in an *in vitro* model of ARDS

BAL from patients with sepsis-related ARDS enrolled to this study was rendered acellular by centrifugation at 500g. The acellular BAL was then pooled and mixed in a 1:1 ratio with 10% FCS/RPMI. To elicit functional changes associated with ARDS, AMs were treated with this 50% ARDS BAL mixture. AMs were also treated with a 1:1 mixture of 0.9% Saline and 10% FCS/RPMI, as vehicle control (VC). Other treatments given in conjunction with 50% ARDS BAL or saline included 200nM Y-27632 dihydrochloride (Rho-associated protein kinase inhibitor, Apexbio, USA), 2μM SF1670 (Phosphatase and tensin homolog inhibitor, Selleckchem, USA), and dimethyl sulfoxide (DMSO, vehicle control for Y-27632 and SF1670, Sigma-Aldrich) at a 1:50,000 dilution. ROCK and PTEN inhibitor treatment doses determined by dose response on untreated AM efferocytosis (supplemental figure 8). Other treatments not combined with 50% ARDS BAL or saline included 50ng/ml IFNγ (Peprotech, UK), 1μg/ml Ultra-Pure LPS (Invitrogen), 40ng/ml IL-4 (Peprotech, UK), and 40ng/ml IL-13, Peprotech, UK). Cytokine concentrations were based on published methods (14). The 1µg/ml dose of LPS was based on the lowest dose required to elicit TNFα production from AMs (supplemental figure 9). Efferocytosis, phagocytosis, apoptosis / viability and RNA extraction for gene expression were performed 24 hours after treatment with 50% ARDS BAL. Phenotyping was performed 48 hours after treatment. AM apoptosis and viability were assessed using an apoptosis detection kit (Biolegend) as described below.

Flow cytometric assessment of AM surface markers

AMs were labelled with the following anti-human antibodies or their isotype controls: CD206-APC, CD80-PE, CD163-FITC, Mer-APC, and SIRPα-FITC. Surface marker expression was assessed by an Accuri C6 flow cytometer and software (BD Biosciences). AM population was gated on forward and side-scatter plot. The median fluorescence intensity (MFI) in relevant channels from isotype control AMs was subtracted from the MFIs of stained AMs, to give the net MFI for each antibody fluorophore.

Assessment of AM gene expression

RNA was isolated from AMs using Nucleospin RNA kits (Machery-Nagel) as per manufacturer’s instructions. RNA quantity was assessed with the NanoDrop 2000 UV-Vis Spectrophotometer (ThermoFisher). One-Step Quantifast Probe RT-PCR Kits (Qiagen) were used to assess gene expression with a CFX384 Touch Real-Time PCR Detection System (BioRad). Taqman^®^ gene expression assays (ThermoFisher) were purchased for 18S on VIC-MGB (ref 4318839) and RAC1 on FAM-MGB (Hs01025984_m1). PCR conditions were used as per manufacturer’s recommendation. Triplicate data was analysed using CFX Maestro software (BioRad). Relative quantification of target gene mRNA was calculated relative to expression of 18s endogenous control gene.

Assessment of neutrophil apoptosis

Flow cytometric assessment of neutrophil apoptosis was performed using a Fluorescein Isothiocyanate (FITC)-conjugated Annexin V and 7-aminoactinomycin D (7-AAD) apoptosis detection kit (Biolegend): As per kit instructions, cells that were Annexin V only positive were classified as early apoptotic, whereas cells which were doubly positive for Annexin V and 7-AAD were classified as late apoptotic / necrotic. Morphological analysis of BAL cytospins by 2 assessors blinded to sample origin was used as an additional method of assessing neutrophil apoptosis. Morphological changes of apoptosis include nuclear fragmentation, chromatin aggregation and cytoplasmic vacuolation(15).

Quantification of inflammatory mediators and endotoxin

BAL inflammatory cytokines (Interleukin-1β/1ra/6/8/10, tumour necrosis factor-α, macrophage chemoattractant protein-1, vascular endothelial growth factor and interferon-γ) were measured by a commercially available custom Magnetic Luminex® Performance Assay (R&D Systems, UK) as per manufacturer’s instructions. BAL endotoxin content was quantified by use of a Pierce Limulus Amebocyte Lysate chromogenic endotoxin quantification kit (ThermoFisher Scientific, U.K) as per manufacturer’s instructions. For each molecular analysis, all samples were processed and assayed simultaneously.

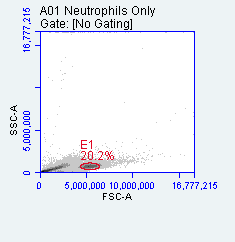

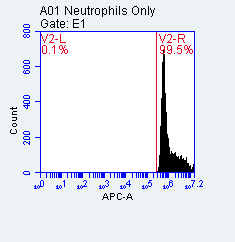

**A**

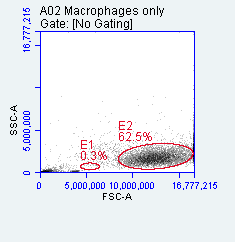

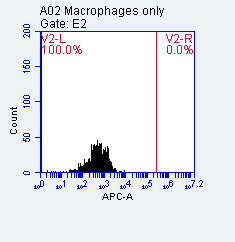

**B**

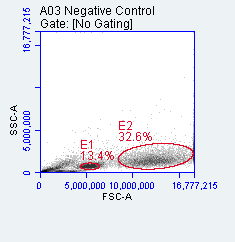

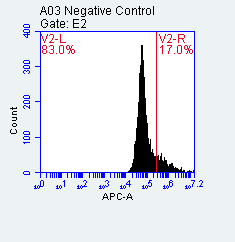

**C**

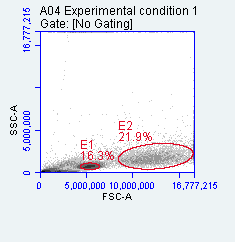

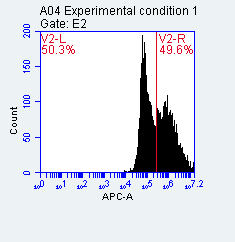

**D**

**Supplemental Figure 1: Example flow plots and histograms for efferocytosis assay**

Forward vs side scatter plots and fluorescence histograms of control and test samples used to set up efferocytosis assay. Alveolar macrophages (AMs) used in this example are from the lung resection of a never smoking patient. Doublet discrimination was also performed on all samples but not shown. All fluorescence histograms are of the APC (FL4) channel, which corresponds to the emission spectra of CellTracker^TM^ Deep Red. **A:** Stained apoptotic neutrophils were used to gate neutrophils (E1) on forward vs side scatter plot, and as a positive control used to determine the positive threshold on the APC fluorescence histogram. **B:** Tube containing AMs only used to gate macrophages on forward vs side scatter plot (E2), and used to confirm that AMs alone do not fluoresce above the positive threshold in the APC channel. **C:** Negative control (Cytochalasin D treated AMs incubated with neutrophils) in which efferocytosis has been inhibited. The APC fluorescence histogram has been gated on E2 (macrophage gate). Since efferocytosis has been inhibited, any fluorescence in the APC channel detected above the positive threshold is due to neutrophil adherence to the surface of AMs, and not engulfment. This background fluorescence value is 17%. **D:** Experimental condition (untreated AMs incubated with neutrophils). The APC fluorescence histogram has been gated on E2 (AM gate), and shows that 49.6% of AMs are APC-positive. However, the background fluorescence value of 17% from the negative control must be subtracted to determine the true efferocytosis index. The efferocytosis index for this sample would therefore be calculated as 32.6%.

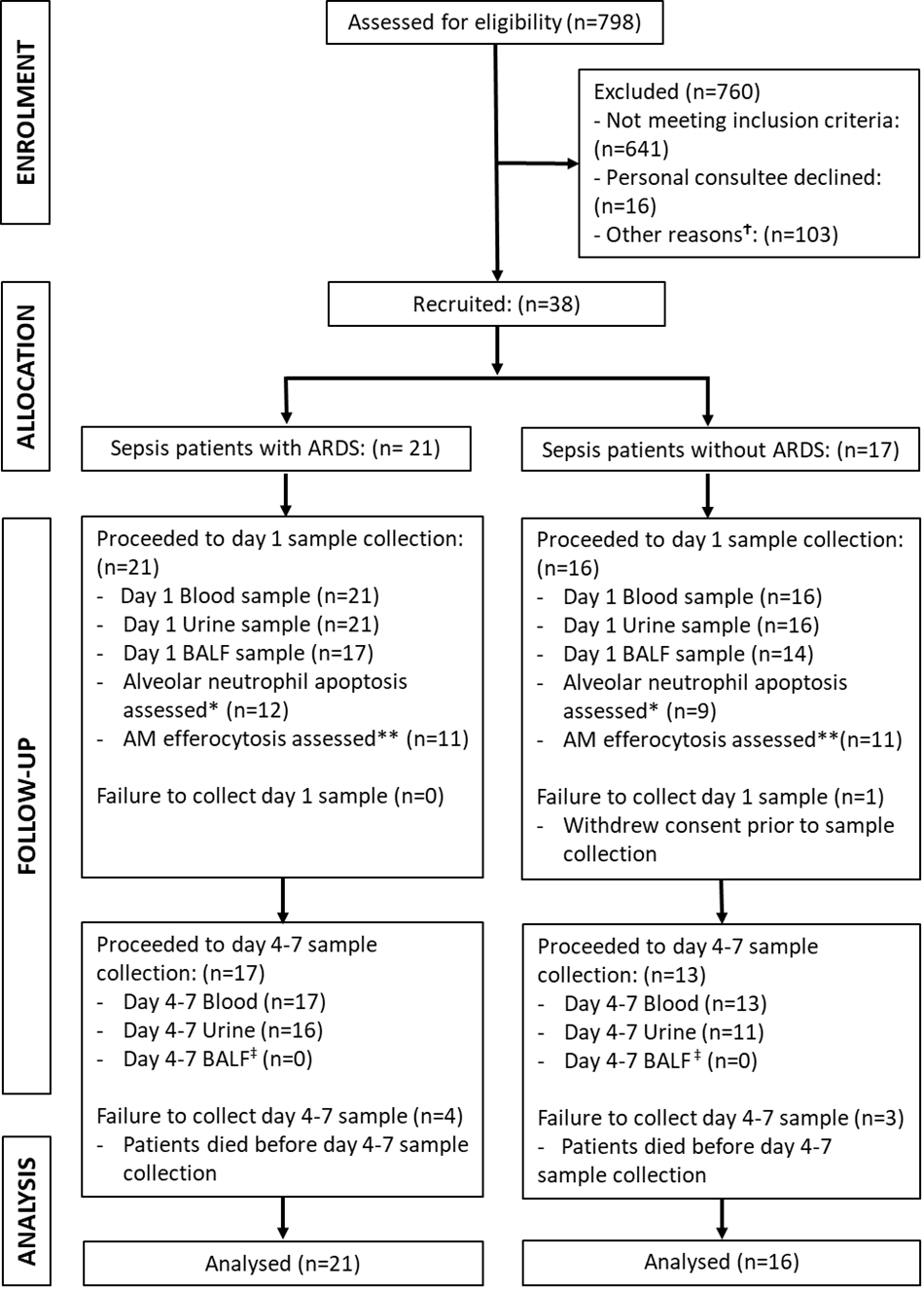

**Supplemental Figure 2: Flow diagram for AM-ARDS study.**

BAL = Broncho-alveolar lavage. AM = alveolar macrophage. **^Ϯ^**The predominant ‘other reason’ for excluding patients was if the ICU consultant responsible for their clinical care did not approve a bronchoscopy. *****Neutrophil apoptosis was not initially assessed and was only added to the study protocol after recruitment had already begun. ******The average AM yield was 1 million per patient. Therefore, it was often difficult to perform both efferocytosis and phagocytosis assays. The efferocytosis assay was given priority; if there were AMs remaining after allocation for efferocytosis assay, then phagocytosis assays would be performed. AM phagocytosis assays were performed on AMs from 3 sepsis patients with ARDS, and 8 sepsis patients without ARDS. **^‡^**A second BAL sample was not obtained from any of the patients, following discussion with the ICU consultant in charge of the patients’ clinical care. There was thought to be no clinical benefit and potential unnecessary risk to repeat bronchoscopy in these patients once a BAL microbiology sample had already been obtained during day 1 sampling.

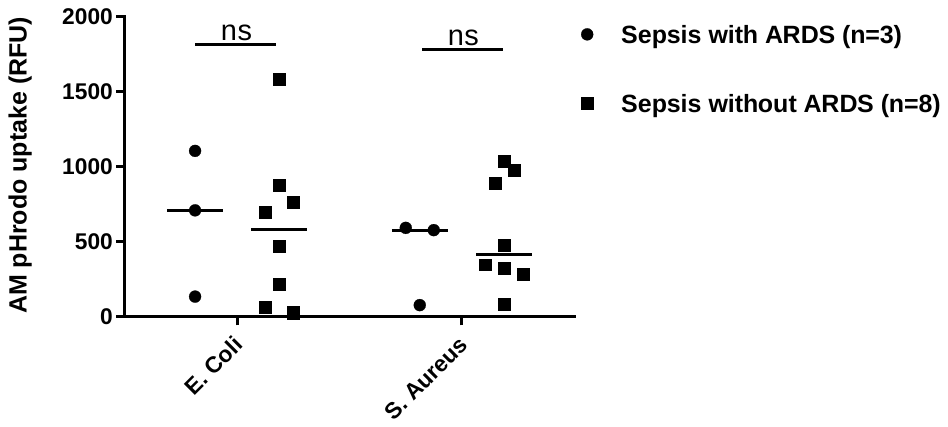

**Supplemental Figure 3: Alveolar macrophage phagocytosis in sepsis patients with and without ARDS**

Alveolar macrophage (AM) phagocytosis assessed by pHrodo bioparticle uptake in sepsis patients with or without ARDS. Error bars shown as medians, statistical analysis by Mann Whitney Test. No significant difference observed between AM phagocytic index between sepsis patients with and without ARDS. This was observed with both *E. coli* (medians 707 vs 580, p = 0.776), and *S. Aureus* (576 vs 411, p = 0.776) pHrodo® bio-particles.

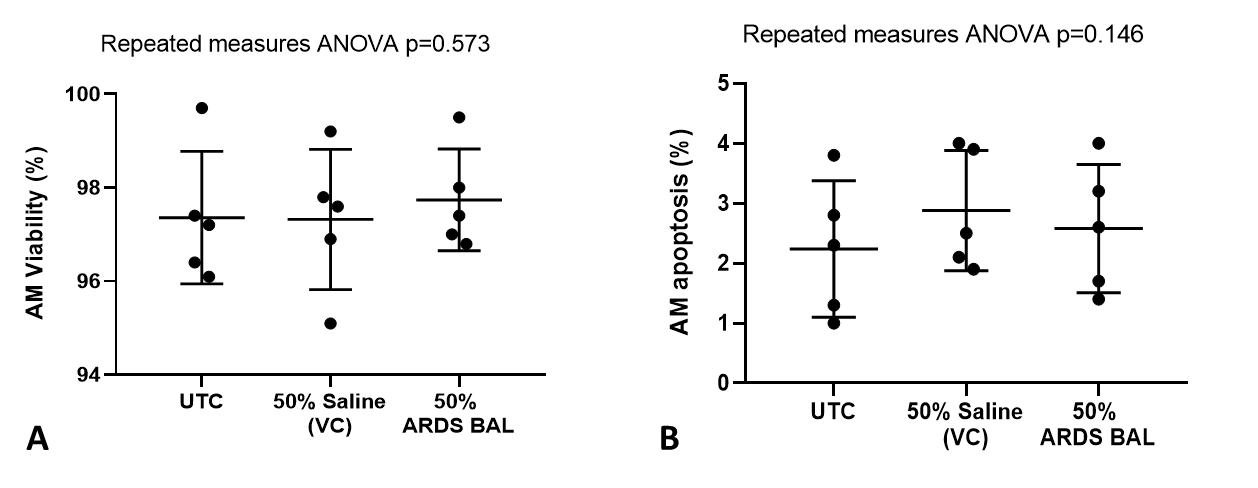

**Supplemental Figure 4: Effect of ARDS BAL treatment on alveolar macrophage viability and apoptosis.**

UTC = Untreated control (RPMI + 10% FBS). VC = Vehicle control (50% Saline). Data shown as mean and standard deviation, n=5 for each group. **A:** VC treatment and 50% ARDS BAL treatment have no significant effect on AM viability (repeated measures ANOVA p=0.57). **B:** VC treatment and 50% ARDS BAL treatment have no significant effect on AM apoptosis (repeated measures ANOVA p=0.15).

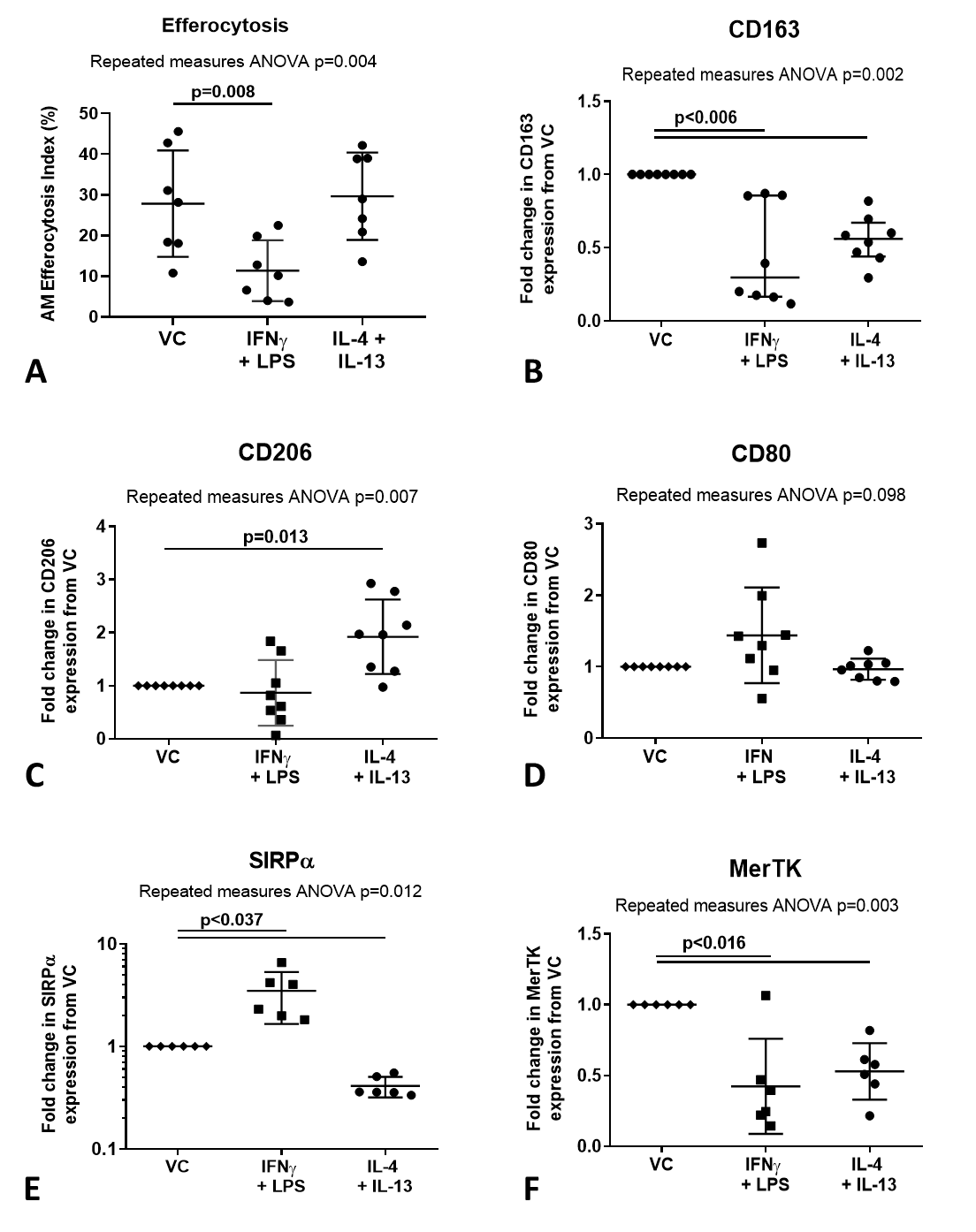

**Supplemental Figure 5: Effect of pro- and anti-inflammatory mediators on alveolar macrophage efferocytosis and surface receptor expression**

VC = Vehicle Control (distilled water added at 1:500 to RPMI + 10% FBS). IL-4 + IL-13 = interleukins 4 & 13 used at 40ng/ml each. IFNγ = 50ng/ml interferon γ. LPS = 1μg/ml lipopolysaccharide. **A:** Data shown as mean and standard deviation, n=7 for all groups. Cytokine treatment significantly affected AM efferocytosis (repeated measures ANOVA p=0.004). IFNγ and LPS treatment significantly reduced AM efferocytosis compared to VC (Dunnett’s multiple comparisons test mean difference 16.5%, p=0.008). IL-4 and IL-13 had no significant effect on AM efferocytosis compared to VC (Dunnett’s multiple comparisons test mean difference -1.8%, p=0.897). **B-F**: Statistical analysis by repeated measures ANOVA with Dunnett’s multiple comparison test, n=6-8. Linear y-axis used for all graphs, except SIRPα (**D**) for which a log scale y-axis was used. **B:** Cytokine treatments significantly affected AM surface expression of CD163 (repeated measures ANOVA p=0.002). Compared to treatment with VC, AM surface expression of CD163 was significantly decreased following treatment with IFNγ + LPS (mean of differences -0.55, p=0.005), and IL-4 + IL-13 (mean of differences -0.45, p=0.002). **C:** Cytokine treatments significantly affected AM surface expression of CD206 (repeated measures ANOVA p=0.007). Compared to treatment with VC, AM surface expression of CD206 was significantly increased following treatment with IL-4 + IL-13 (mean of differences 0.92, p=0.013). There were no significant changes in CD206 expression following treatment with IFNγ + LPS (mean of differences -0.13, p=0.777). **D:** Cytokine treatments did not significantly affect AM surface expression of CD80 (repeated measures ANOVA p=0.098). **E:** Cytokine treatments significantly affected AM surface expression of SIRPα (repeated measures ANOVA p=0.012). Compared to treatment with VC, AM surface expression of SIRPα was significantly increased following treatment with IFNγ + LPS (median of differences 2.48, p=0.036). Compared to treatment with VC, AM surface expression of SIRPα was significantly decreased following treatment with IL-4 + IL-13 (median of differences -0.59, p<0.0001). **F:** Cytokine treatments significantly affected AM surface expression of MerTK (repeated measures ANOVA p=0.003). Compared to treatment with VC, AM surface expression of MerTK was significantly decreased following treatment with IFNγ + LPS (mean of differences -0.58, p=0.015), and IL-4 + IL-13 (mean of differences -0.47, p=0.004.

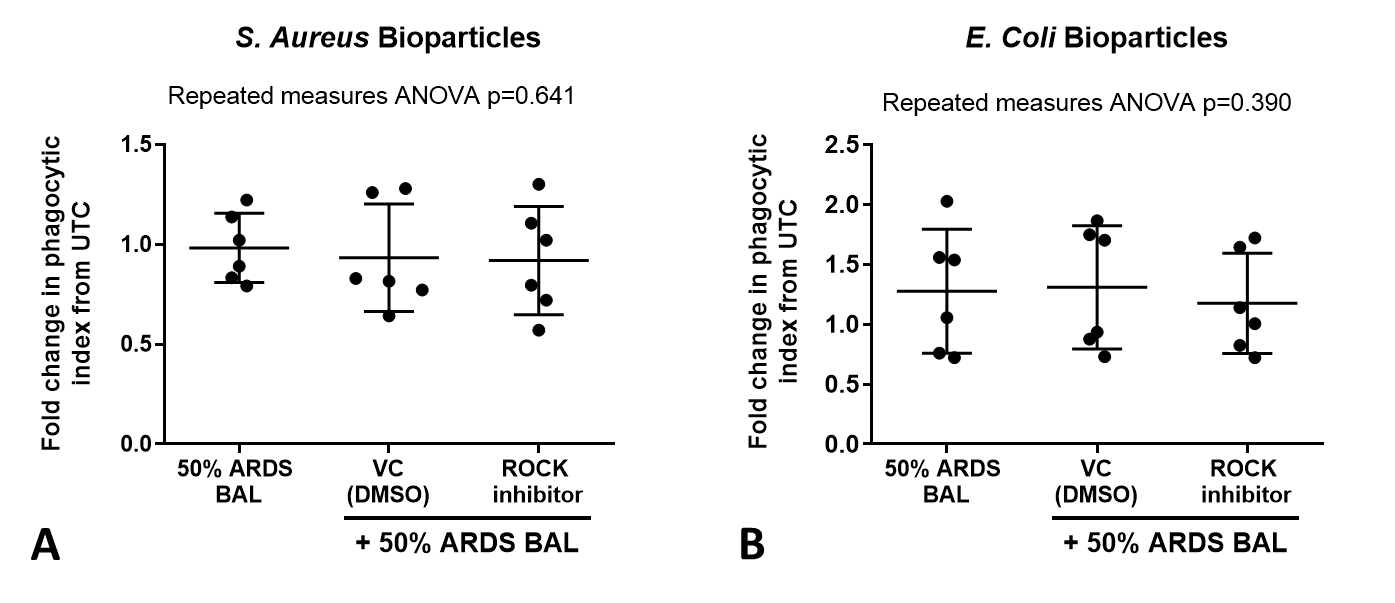

**Supplemental Figure 6: Effect of ARDS BAL and ROCK-inhibitor treatment on alveolar macrophage phagocytosis**

UTC = Untreated control. VC = Vehicle Control for ROCK-inhibitor (Dimethyl Sulfoxide [DMSO] at 1:50,000 dilution). ROCK-inhibitor = 200nM Y-27632 dihydrochloride; Rho-associated protein kinase inhibitor. 50% Saline acted as vehicle control for 50% ARDS BAL treatment. Data shown as mean and standard deviation, corrected to fold change in phagocytic index from UTC. Statistical analysis by repeated measures ANOVA. **A:** Addition of VC or ROCK-inhibitor to 50% ARDS BAL mixture had no significant effect on AM phagocytosis of *S. Aureus* pHrodo® bioparticles (repeated measures ANOVA p=0.641). **B:** Addition of VC or ROCK-inhibitor to 50% ARDS BAL mixture had no significant effect on AM phagocytosis of *E. coli* pHrodo® bioparticles (repeated measures ANOVA p=0.390).

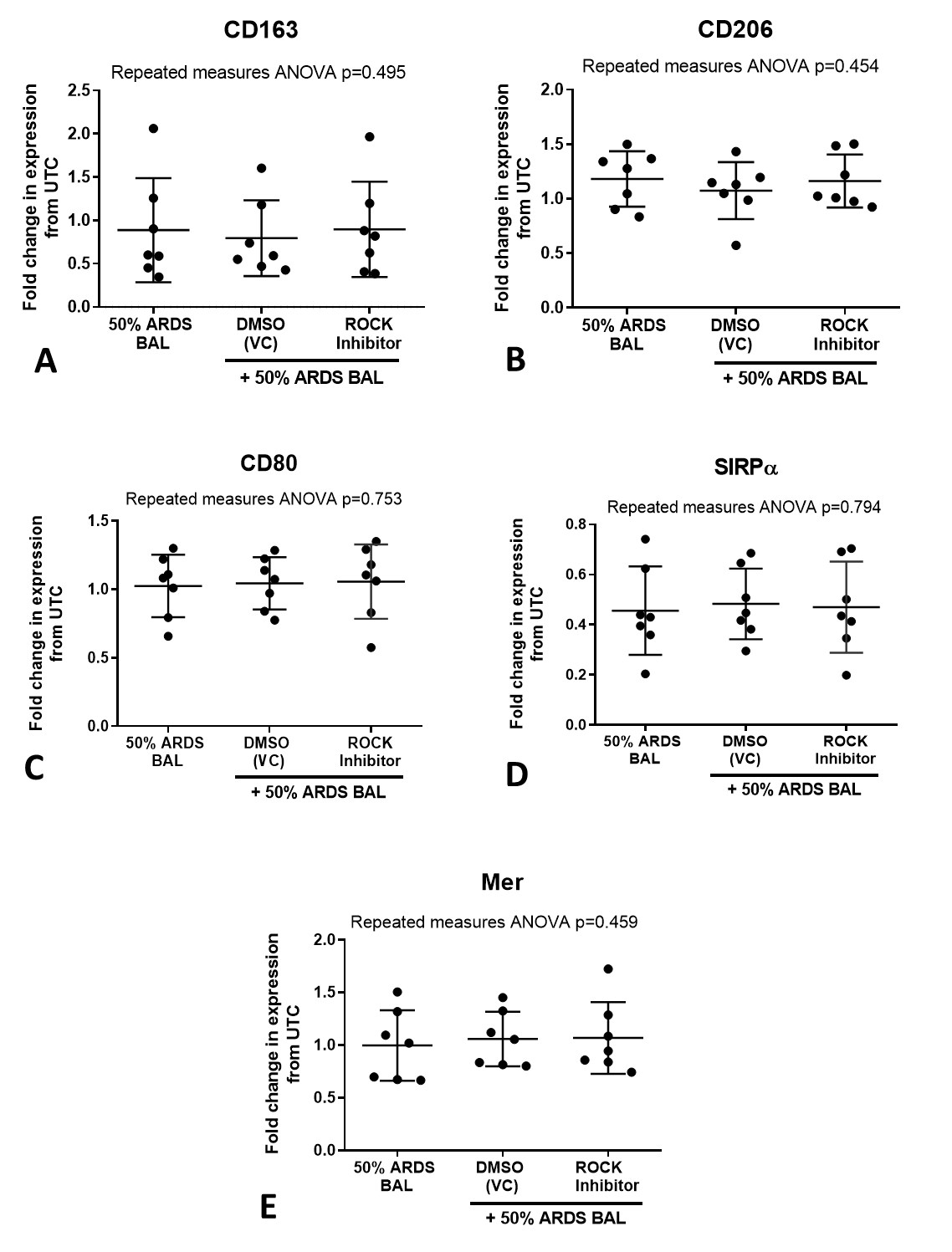

**Supplemental Figure 7: Effect of ROCK-inhibitor on alveolar macrophage phenotype and surface receptor expression**

VC = Vehicle Control (Dimethyl Sulfoxide [DMSO] at 1:50,000 dilution). ROCK-inhibitor = 200nM Y-27632 dihydrochloride; Rho-associated protein kinase inhibitor. Mer = Mer receptor tyrosine kinase. SIRPα = Signal regulatory protein alpha. Statistical analysis by paired t-test, n=7 for all groups. **A-E:** Addition of ROCK-inhibitor to 50% ARDS BAL treatment had no significant effect on AM surface expression of CD163, CD206, CD80, SIRPα and MerTK, compared to treatment with VC + ARDS BAL (p>0.05 for all) or ARDS BAL alone (repeated measures ANOVA p>0.45 for all).

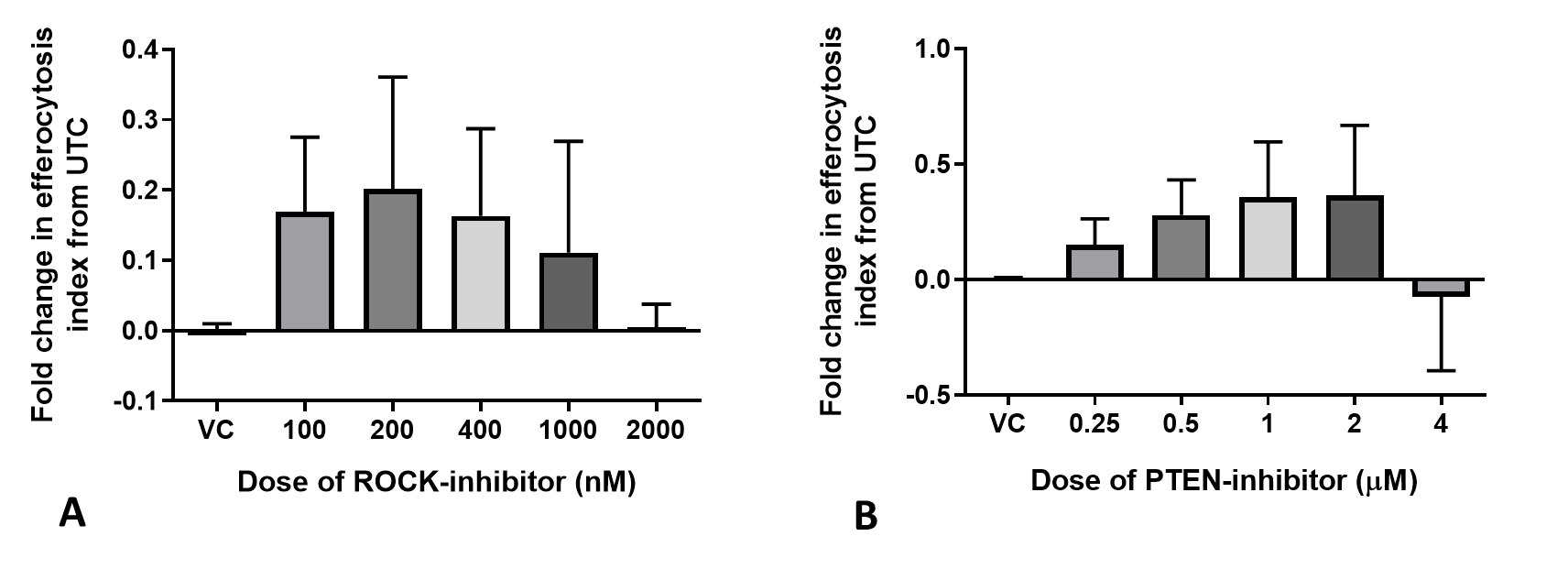

**Supplemental Figure 8: Dose response of ROCK-inhibitor and PTEN-inhibitor on the efferocytosis index of uninjured AMs.**

Data shown as mean and standard deviation, n=4 all groups. UTC = Untreated control. VC = Vehicle Control (Dimethyl Sulfoxide [DMSO] at 1:50,000 dilution). ROCK-inhibitor = 200nM Y-27632 dihydrochloride; Rho-associated protein kinase inhibitor. PTEN-inhibitor = 2μM SF1670; Phosphatase and tensin homolog inhibitor. **A:** The dose of ROCK-inhibitor which elicited the greatest fold change in the efferocytosis index of uninjured alveolar macrophages was 200mM, equal to the IC_50_ (50% of the maximal inhibitory concentration) **B:** The dose of PTEN-inhibitor which elicited the greatest fold change in the efferocytosis index of uninjured alveolar macrophages was 2µM, equal to IC_50_.

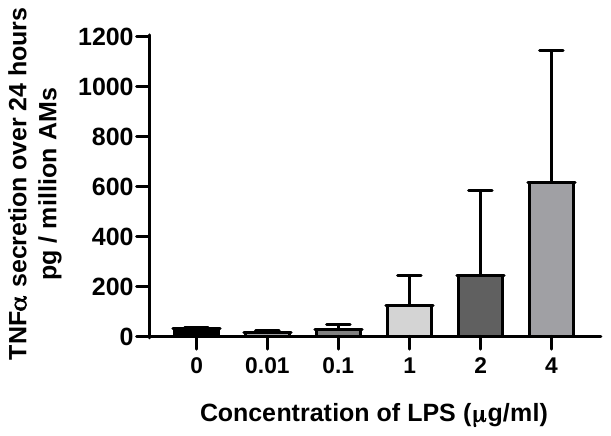

**Supplemental Figure 9: Dose response of LPS on alveolar macrophage secretion of TNF-α**

LPS = Lipopolysaccaride. TNF-α = Tumour necrosis factor-α. UTC = Untreated control. n=3. LPS only stimulated AM secretion of TNFα at doses ≥ 1µg/ml.

|  | **Sepsis patients with ARDS (n=17)** | **Sepsis patients without ARDS (n=14)** | **Mann Whitney Test** |
| --- | --- | --- | --- |
| **IL-6**  **(pg/ml)** | 74.6  (25.9 – 724.4) | 84.0  (9.2 – 115.4) | p = 0.401 |
| **IL-8**  **(pg/ml)** | 4163  (652 – 18432) | 248  (109 – 2842) | **p = 0.019** |
| **IL-1ra**  **(pg/ml)** | 4023  (1067– 10980) | 834  (289 – 7329) | p = 0.110 |
| **IL-1β**  **(pg/ml)** | 46.7  (7.8 – 471.5) | 9.4  (7.7 – 82.4) | p = 0.331 |
| **TNFα**  **(pg/ml)** | 6.7  (0 - 22.6) | 0  (0 – 8.9) | p = 0.153 |
| **IL-10**  **(pg/ml)** | 5.6  ( 5.6 – 9.0) | 5.6  (5.6 – 5.6) | p = 0.404 |
| **VEGF**  **(pg/ml)** | 208.2  (84.7 – 441.0) | 109.4  (16.6 – 354.5) | p = 0.173 |
| **MCP-1**  **(pg/ml)** | 1048  (499 – 3175) | 253  (101 – 1359) | **p = 0.012** |

**Supplemental Table 1: BAL cytokines from sepsis patients with and without ARDS on day 1 of recruitment**

All data presented as median and interquartile range. IL = interleukin. TNFα = tumour necrosis factor α. VEGF = vascular endothelial growth factor. MCP-1 = monocyte chemoattractant protein-1.

| **Characterization of pooled ARDS patient BAL** | |
| --- | --- |
| **IL-6** | 453 pg/ml |
| **IL-8** | 4268 pg/ml |
| **IL-1β** | 98 pg/ml |
| **IL-1ra** | 3023 pg/ml |
| **IL-10** | 6 pg/ml |
| **TNF-α** | 3 pg/ml |
| **VEGF** | 209 pg/ml |
| **MCP-1** | 1045 pg/ml |
| **IFN-γ** | 0 pg/ml |
| **LPS** | 57 pg/ml |

**Supplemental Table 2: Characterization of pooled ARDS patient BAL**

IFN-γ = Interferon-γ. IL = Interleukin. TNF-α = Tumor necrosis factor-α. MCP-1 = macrophage chemoattractant protein-1. LPS = Lipopolysaccharide. VEGF = vascular endothelial growth factor.
